# Epidemiology and role of SARS-CoV-2 Linkage in Paediatric Inflammatory Multisystem Syndrome (PIMS): A Canadian Paediatric Surveillance Program National Prospective Study

**DOI:** 10.1101/2022.05.27.22275613

**Authors:** Tala El Tal, Marie-Paule Morin, Shaun K. Morris, Daniel S. Farrar, Roberta A Berard, Fatima Kakkar, Charlotte Moore Hepburn, Krista Baerg, Camille Beaufils, Terri-Lyn Bennett, Susanne M Benseler, Guillaume Beaudoin-Bussières, Kevin Chan, Claude Cyr, Nagib Dahdah, Elizabeth J. Donner, Olivier Drouin, Rojiemiahd Edjoc, Maryem Eljaouhari, Joanne E. Embree, Catherine Farrell, Andrés Finzi, Sarah Forgie, Ryan Giroux, Kristopher T. Kang, Melanie King, Melanie Laffin Thibodeau, Bianca Lang, Ronald M. Laxer, Thuy Mai Luu, Brian W. McCrindle, Julia Orkin, Jesse Papenburg, Catherine M. Pound, Victoria E. Price, Jean-Philippe Proulx-Gauthier, Rupeena Purewal, Manish Sadarangani, Marina l. Salvadori, Roseline Thibeault, Karina A. Top, Isabelle Viel-Thériault, Elie Haddad, Rosie Scuccimarri, Rae S. M. Yeung

**Affiliations:** Division of Rheumatology, Department of Pediatrics, The Hospital for Sick Children, University of Toronto, Ont., Canada; Division of Paediatric Rheumatology-Immunology, CHU Sainte-Justine, Department of Pediatrics, University of Montreal, Montreal, Que., Canada; Department of Pediatrics, Faculty of Medicine, University of Toronto; Division of Infectious Diseases, The Hospital for Sick Children, Toronto, Ont., Canada; Centre for Global Child Health and Child Health Evaluative Sciences, The Hospital for Sick Children; Clinical Public Health, Dalla Lana School of Public Health, University of Toronto, Toronto, Ont., Canada; Division of Rheumatology, Department of Pediatrics, Children’s Hospital, London Health Sciences Centre, London, Ont., Canada; Division of Infectious Diseases, CHU Sainte-Justine, Montreal, Que., Canada; Department of Pediatrics, Division of Paediatric Medicine, Hospital for Sick Children, University of Toronto, Toronto, Ont., Canada; Department of Pediatrics, University of Saskatchewan, Saskatoon, Sask., Canada; Public Health Agency of Canada, Ottawa, Canada; Alberta Children’s Hospital Research Institute, Cumming School of Medicine, University of Calgary, Calgary, AB, Canada; Division of Rheumatology, Department of Pediatrics, Alberta Children’s Hospital, University of Calgary, Calgary, AB, Canada; Department of microbiology, immunology and infectious diseases, Université de Montréal, QC, Canada; Centre de Recherche du CHUM, Montreal, QC, Canada; Department of Children’s and Women’s Health, Trillium Health Partners, Mississauga, Ont., Canada; Service de Soins Intensifs Pédiatriques, Centre Hospitalier Universitaire de Sherbrooke, Sherbrooke, Que., Canada; Division of Pediatric Cardiology, CHU Sainte-Justine, Department of Pediatrics, University of Montreal, Montreal, Que., Canada; Division of Neurology, The Hospital for Sick Children, University of Toronto, Toronto, Ontario, Canada; Division of General Pediatrics, Department of Pediatrics, CHU Sainte-Justine, Montréal, Que; Department of Social and Preventive Medicine, School of Public Health, Université de Montréal, Montréal, Que., Canada; Department of Paediatrics and Child Health, University of Manitoba, Winnipeg, Manitoba, Canada; Department of Medical Microbiology and Infectious Diseases, University of Manitoba, Winnipeg, Manitoba, Canada; Division of Paediatric Intensive Care, Department of Pediatrics, CHU Sainte-Justine, Montréal, Que., Canada; Division of Infectious Diseases, Department of Pediatrics, University of Alberta, Edmonton, Alta., Stollery Children’s Hospital, Edmonton, Alta., Canada; Women’s and Children’s Health Program, St. Michael’s Hospital, Unity Health Toronto, Toronto, Ontario; Department of Pediatrics, University of British Columbia, Vancouver, BC, Canada; Canadian Paediatric Surveillance Program, Canadian Paediatric Society, Ottawa, Canada; Division of Rheumatology, Department of Pediatrics, Dalhousie University, Halifax, NS, Canada; The Labatt Family Heart Centre, The Hospital for Sick Children, Department of Pediatrics, University of Toronto, Toronto, Ont., Canada; Child Health Evaluative Sciences, The Hospital for Sick Children, Toronto, Ont., Canada; Division of Pediatric Infectious Diseases, Department of Pediatrics, Montreal Children’s Hospital, Montréal, Que., Canada; Division of Microbiology, Department of Clinical Laboratory Medicine, McGill University Health Centre, Montréal, Que., Canada; Division of Consulting Pediatrics, Department of Pediatrics, Children’s Hospital of Eastern Ontario, Ottawa, Ont, Canada; Division of Paediatric Hematology/Oncology, Department of Pediatrics, Dalhousie University, Halifax, NS., Canada; Department of Pediatrics, CHU de Québec-Université Laval, Québec City, Quebec, Canada; Division of Paediatric Infectious Diseases, Jim Pattison Children’s Hospital, Saskatchewan Health Authority, Saskatoon, Sask., Canada; Vaccine Evaluation Center, BC Children’s Hospital Research Institute, Vancouver, BC., Canada; Division of Pediatric Infectious Diseases, Department of Paediatrics, CHU de Quebec-University of Laval, Quebec City, Que., Canada; Department of Pediatrics, Dalhousie University, Halifax, NS. Canada; Division of Infectious Diseases, Department of Pediatrics, CHU de Québec-Université Laval, Québec City, Que., Canada; Division of Paediatric Rheumatology, Montreal Children’s Hospital/McGill University Health Centre, Montreal, Quebec, Canada; Cell Biology Program, The Hospital for Sick Children Research Institute, Toronto, Ontario, Canada; Department of Immunology and Institute of Medical Science, University of Toronto, Canada

## Abstract

**Background:** Paediatric inflammatory multisystem syndrome (PIMS) is a rare but serious condition temporally associated with SARS-CoV-2 infection. Using the Canadian Paediatric Surveillance Program (CPSP), a national surveillance system, we aimed to 1) study the impact of SARS-CoV-2 linkage on clinical and laboratory characteristics, and outcomes in hospitalized children with PIMS across Canada 2) identify risk factors for ICU admission, and 3) establish the minimum national incidence of hospitalizations due to PIMS and compare it to acute COVID-19.

**Methods:** Weekly online case reporting was distributed to the CPSP network of more than 2800 pediatricians, from March 2020 to May 2021. Comparisons were made between cases with respect to SARS-CoV-2 linkage. Multivariable modified Poisson regression was used to identify risk factors for ICU admission and Minimum incidence proportions were calculated.

**Findings:** In total, 406 PIMS cases were analyzed, of whom 202 (49· 8%) had a positive SARS-CoV-2 linkage, 106 (26· 1%) had a negative linkage, and 98 (24· 1%) had an unknown linkage. The median age was 5· 4 years (IQR 2· 5–9· 8), 60% were male, and 83% had no identified comorbidities. Compared to cases with a negative SARS-CoV-2 linkage, children with a positive SARS-CoV-2 linkage were older (8· 1 years [IQR 4· 2–11· 9] vs. 4· 1 years [IQR 1· 7–7· 7]; p<0· 001), had more cardiac involvement (58· 8% vs. 37· 4%; p<0· 001), gastrointestinal symptoms (88· 6% vs. 63· 2%; p<0· 001), and shock (60· 9% vs. 16· 0%; p<0· 001). At–risk groups for ICU admission include children ≥ 6 years and those with a positive SARS-CoV-2 linkage. No deaths were reported. The minimum incidence of PIMS hospitalizations during the study period was 5· 6 hospitalizations per 100,000 population <18 years.

**Interpretation:** While PIMS is rare, almost 1 in 3 hospitalized children required ICU admission and respiratory/hemodynamic support, particularly those ≥ 6 years and with a positive SARS-CoV-2 linkage.

**Funding:** Financial support for the CPSP was received from the Public Health Agency of Canada.

## Introduction

In April 2020, international reports simultaneously began describing children presenting with a rare but serious multisystem inflammatory condition, temporally associated with an antecedent SARS-CoV-2 infection.^1,2,3^ Clinical features included fever and hyperinflammation, Kawasaki Disease (KD)-like presentation, and shock-like state partially overlapping with toxic shock syndrome (TSS).^4,5^ Several similar but not identical case definitions were developed by different international organizations, including paediatric inflammatory multisystem syndrome temporally associated with SARS-CoV-2 (PIMS-TS) and multisystem inflammatory syndrome in children (MIS-C).^6,7,8^ The Canadian Paediatric Surveillance Program (CPSP) is a national voluntary surveillance system jointly operated by the Canadian Paediatric Society and Public Health Agency of Canada (PHAC) that typically gathers information about rare diseases and conditions of high morbidity and mortality despite low frequency.^9^ In March 2020, it launched a national study of acute SARS-CoV-2 infection. In May 2020, recognizing the emergence of a post-infectious hyperinflammatory syndrome associated with this novel virus, the CPSP study was rapidly adapted to include surveillance of this syndrome.^10,11^

Using the reported information, we aimed to 1) study the impact and clinical implications of requisite SARS-CoV-2 linkage on clinical and laboratory features, and outcomes, of hospitalized children presenting with this syndrome, 2) gain a better understanding of risk factors associated with ICU admission, and 3) define the minimum incidence of hospitalizations due to this syndrome and compare it to acute COVID-19.

## Methods

### Study design and participants

In the absence of a clear and universally accepted case definition in early 2020, and recognizing the need for rapid launch of surveillance, the CPSP developed a surveillance case definition of suspected paediatric inflammatory multisystem syndrome (PIMS) as best understood at the time, consisting of (1) persistent fever (>38.0 °C for ≥ 3 days), (2) elevated inflammatory markers (C-reactive protein [CRP], erythrocyte sedimentation rate [ESR], or ferritin), (3) features of KD and/or TSS, and (4) ensuring no alterative etiology to explain the clinical presentation.^11^ Of note, this case definition was originally intended as a surveillance mechanism that helped capture trends, and was not made for the purpose of clinical diagnosis. In addition, careful consideration was made about the addition of SARS-CoV-2 testing, electing to keep this optional, in the face of limited testing capacity across Canada at the onset of the pandemic, and public health reporting capacity.

Through the CPSP platform, over 2,800 paediatricians and paediatric sub-specialists were surveyed weekly from March 2020 to May 31, 2021 to report all hospitalized patients <18 years of age meeting the PIMS surveillance case definition. The full protocol and case report form for the study can be found at: https://www.cpsp.cps.ca/surveillance/study-etude/covid-19. ^11^ Cases of “PIMS” that were included in the primary analysis were those who met study definition for PIMS. A post hoc definition for MIS-C using the subsequently developed WHO case definition, was also applied on these retained cases [7]. Results of this are described in Appendix 1. Exclusion criteria included cases that did not meet PIMS criteria upon review, incomplete reports, and duplicates. Duplicates were identified using date of birth and other unique data points. Reporting physicians consented to follow-up for 97· 5% of cases to clarify discrepant, missing, or pending data points. The CPSP operates under legal authority derived from Section 4 of the Department of Health Act and Section 3 of the PHAC Act. Research ethics approval was obtained at Health Canada-PHAC (REB #2020-002P), The Hospital for Sick Children (REB #1000070001), the Centre Hospitalier Universitaire Sainte-Justine (IRB #MP-21-2021-2901), and at individual sites as required by local policies. In Quebec, the study was conducted as a multicenter study with clinical data collected by study co-investigators, and serologies from CHUSJ done for research purposes (REB #3195) prior to introduction of clinical testing were also included.

### Demographic, clinical and laboratory data

Key demographic, clinical, laboratory, and treatment data were captured.^11^ Coronary artery abnormalities were defined by lesions Z-scores ≥ 2 SD which has been further disaggregated into two categories of <2· 5 and ≥ 2· 5 SD with body surface area normalized measurements.^12^ The phases of the COVID-19 pandemic have been defined as “first wave” (March–August 2020) and “second and third” waves combined (September 2020–May 2021).

### Study definitions

“PIMS” is defined as all cases that met the definition for PIMS and/or MIS-C (complete algorithms described in Appendix 1). These cases were further sub-divided into three groups 1) “PIMS cases with positive SARS-CoV-2 linkage”, indicates a PIMS case with a positive PCR or rapid test, serology, or close contact, 2) “PIMS cases with negative SARS-CoV-2 linkage”, indicates a PIMS case with a negative serologic test with no other positive SARS-CoV-2 test or close contact, and 3) “PIMS cases with unknown SARS-CoV-2 linkage”, indicates a PIMS case for whom SARS-CoV-2 testing, including serology, was either not conducted or not reported. Reporting physicians were re-contacted throughout the study to ascertain serology results not available at the time of original case reporting and cases were reassigned as appropriate.

In all cases where physicians reported a concurrent microbiologically confirmed or clinically diagnosed infection, at least one infectious disease specialist (authors SKM or FK) and at least one rheumatologist (authors MPM or RB) systematically reviewed all available clinical information to assess the contribution of the concurrent infection to the patient’s clinical presentation. Cases of disagreement were resolved by consensus of all four physicians. Reports of all radiologic findings were also reviewed and categorized as either abnormal, non-specific, or normal.

### Statistical Analysis

Characteristics were described using medians, interquartile ranges, frequencies and percentages. Due to CPSP privacy policies, frequencies between one and four were masked as ‘<5’ and some frequencies were presented as ranges to prevent back-calculation of values <5. To assess differences in SARS-CoV-2 linkage groups, subgroup comparisons were drawn between PIMS cases with positive SARS-CoV-2 linkage and PIMS cases with negative SARS-CoV-2 linkage using χ^2^ tests, Fisher’s exact tests, and Wilcoxon rank-sum tests. To assess differences in continuous laboratory markers, values were log transformed and analyzed using multiple linear regression, adjusting for age category. The temporal lag between weekly PIMS hospitalizations and weekly Canadian SARS-CoV-2 case counts was assessed using Spearman’s rank correlation coefficient.^13^ Multivariable Poisson regression with robust standard errors was conducted to identify within-hospital factors associated with ICU admission due to PIMS, and to avoid inflated interpretations of risk from logistic regression. Secondary analyses were conducted to compare patients meeting versus not meeting criteria for PIMS, by pandemic wave, by age category, and by highest level of care required. A p-value <0· 05 was considered statistically significant for all analyses. Analyses were conducted using Stata version 17· 0.

Finally, we compared the burden of hospitalizations due to PIMS and acute COVID-19 reported to CPSP as follows. Minimum incidence proportions were calculated by dividing the total and age-specific number of hospitalizations, ICU admissions, and deaths by corresponding population denominators retrieved from Statistics Canada.^14^ Confidence intervals were calculated by assuming a Poisson distribution, and were not calculated for values <5. The CPSP PIMS and COVID-19 hospitalization studies were conducted using the same reporting methods and period (i.e., up until May 31, 2021) and thus could be directly compared, however, sensitivity analyses excluding cases from Alberta were conducted as acute COVID-19 data were less complete in that province. Hospitalizations with incidentally identified SARS-CoV-2 infection were excluded from these comparisons, as previously described.^10^

## Results

### Patient demographics

A total of 493 cases were reported to the CPSP PIMS study of whom 406 met the study definition of PIMS (Figure 1) and were hospitalized within the study period. PIMS cases with positive SARS-CoV-2 linkage was reported among 202 children (49· 8%). Of these children, 66· 8% had a positive close contact, 52· 0% had a positive PCR or rapid test, and 48· 0% had positive serology (Table 1, Supplementary Figure 1). Among PCR positive patients, 27· 6% were positive during the hospital stay while the remaining patients had a positive test a median of 4· 6 weeks (IQR 3· 4–5· 9) prior to hospitalization. Among children with reported serologies (n=213), 54· 5% had a negative result (n=116) though ten of these cases had a positive PCR or close contact. Therefore, among all PIMS cases, 26· 1% had negative SARS-CoV-2 linkage (n=106), while 24· 1% had unknown SARS-CoV-2 linkage (n=98), with no serology conducted or reported to CPSP.

**Table 1.** Demographic and outcome characteristics of children hospitalized with PIMS in Canada from March 2020–May 2021, overall and by SARS-CoV-2 linkage group.

| Characteristics | All cases | SARS-CoV-2 linkage<sup>1</sup> |  |  |  |
| --- | --- | --- | --- | --- | --- |
|  |  | Positive | Negative | P value | Unknown |
| **Number of PIMS hospitalizations, N** | 406 | 202 | 106 | --- | 98 |
| **Age (years), median (IQR)** | 5.4 (2.5–9.8) | 8.1 (4.2–11.9) | 4.1 (1.7–7.7) | <0.001 | 3.4 (1.4–6.3) |
| **Age, n (%)** |  |  |  | <0.001 |  |
| <1 year | 43 (10.6) | 9 (4.5) | 15 (14.2) |  | 19 (19.4) |
| 1–5 years | 170 (41.9) | 64 (31.7) | 54 (50.9) |  | 52 (53.1) |
| 6–12 years | 138 (34.0) | 90 (44.6) | 29 (27.4) |  | 19 (19.4) |
| 13–17 years | 55 (13.5) | 39 (19.3) | 8 (7.5) |  | 8 (8.2) |
| **Sex, n (%)** |  |  |  | 0.18 |  |
| Female | 163 (40.1) | 72 (35.6) | 46 (43.4) |  | 45 (45.9) |
| Male | 243 (59.9) | 130 (64.4) | 60 (56.6) |  | 53 (54.1) |
| **Physician-reported population group<sup>2</sup>, n (%)** |  |  |  |  |  |
| White | 157 (38.7) | 64 (31.7) | 47 (44.3) | 0.03 | 46 (46.9) |
| Black | 54 (13.3) | 28 (13.9) | 10 (9.4) | 0.26 | 16 (16.3) |
| East/Southeast Asian | 44 (10.8) | 13 (6.4) | 23 (21.7) | <0.001 | 8 (8.2) |
| South Asian | 43 (10.6) | 26–29 (12.9–14.4) | <5 (<4.7) | 0.01 | 13–16 (13.3–16.3) |
| Arab/West Asian | 28 (6.9) | 14 (6.9) | 9 (8.5) | 0.62 | 5 (5.1) |
| Latin American | 26 (6.4) | 17–20 (8.4–9.9) | 5–8 (4.7–7.5) | 0.23 | <5 (<5.1) |
| Indigenous | 10 (2.5) | 5–8 (2.5–4.0) | <5 (<4.7) | 0.17 | <5 (<5.1) |
| Other | 8 (2.0) | <5 (<2.5) | <5 (<4.7) | 0.42 | <5 (<5.1) |
| Unknown | 69 (17.0) | 45 (22.3) | 16 (15.1) | 0.13 | 8 (8.2) |
| **Positive SARS-CoV-2 linkage, n (%)** | 202 (49.8) | 202 (100.0) | --- | --- | --- |
| Close contact with confirmed SARS-CoV-2 | 135 (33.3) | 135 (66.8) | --- | --- | --- |
| PCR positive | 105 (25.9) | 105 (52.0) | --- | --- | --- |
| Serology positive | 97 (23.9) | 97 (48.0) | --- | --- | --- |
| **History of negative serology, n (%)** | 116 (28.6) | 10 (5.0) | 106 (100.0) | --- | --- |
| **Admitted to ICU, n (%)** | 127 (31.3) | 102 (50.5) | 14 (13.2) | <0.001 | 11 (11.2) |
| **Child died, n (%)** | 0 (0.0) | 0 (0.0) | 0 (0.0) | --- | 0 (0.0) |

ICU = Intensive care unit; IQR = Interquartile range; PCR = Polymerase chain reaction
<sup>1</sup>A positive link indicates a positive PCR or rapid test, serology, or close contact; a negative link indicates a negative serology result with no other positive test or close contact; an unknown link indicates a lack of a positive test or close contact, with serology either not conducted or not reported.
<sup>2</sup>Physicians could report multiple population groups. East/Southeast includes Chinese, Filipino, Japanese, Korean, and Southeast Asian. Indigenous includes First Nations, Inuit, and Métis.

**Figure 1.**
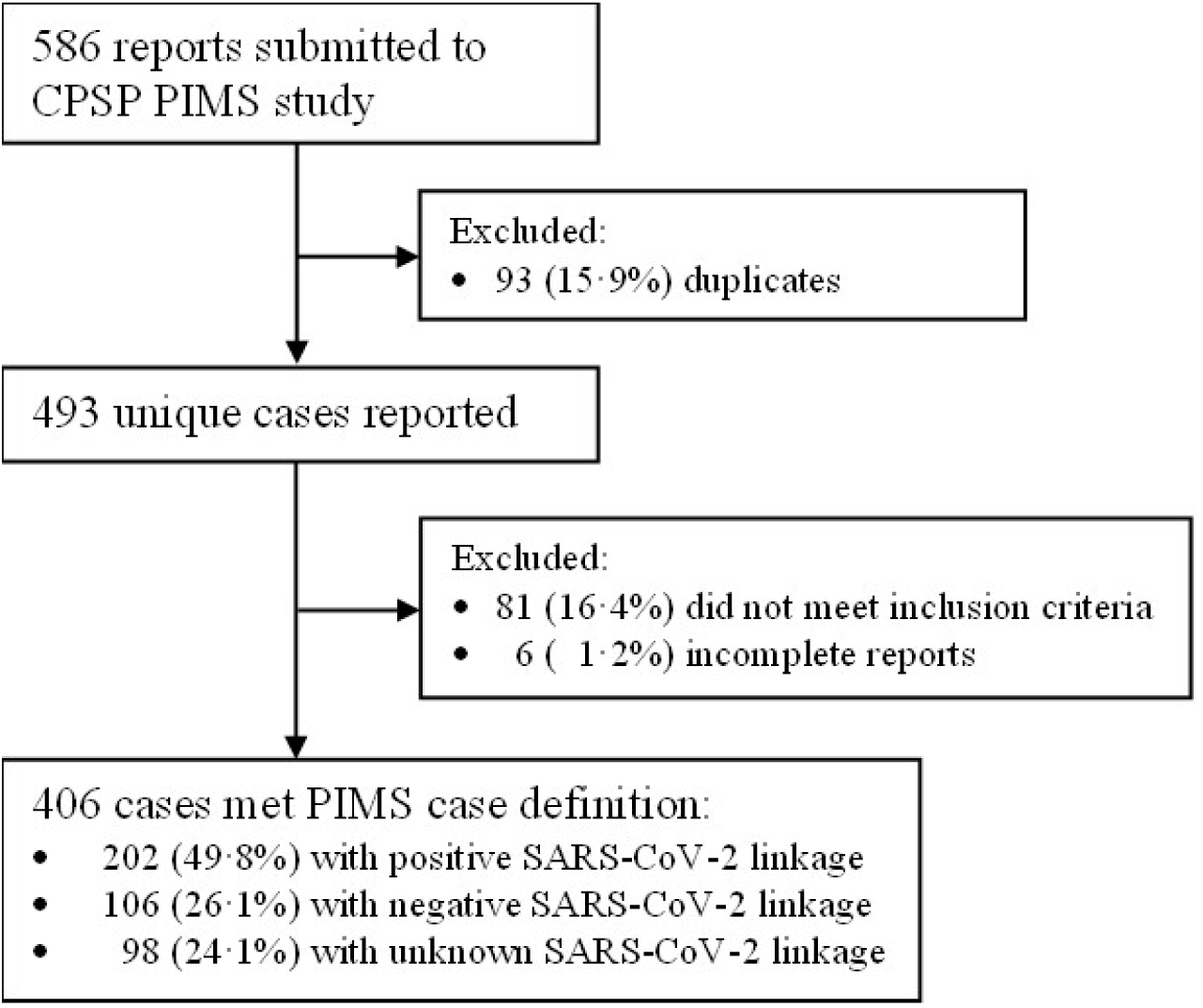
Flow chart of participants meeting the case definitions for PIMS.

The median age among children hospitalized for PIMS was 5· 4 years (IQR 2· 5–9· 8) and more children were male (59· 9%; Table 1). On average, cases with a positive SARS-CoV-2 linkage were significantly older (8· 1 years, IQR 4· 2–11· 9) than PIMS cases with negative SARS-CoV-2 linkage (4· 1 years, IQR 1· 7– 7· 7; p<0· 001), as were cases hospitalized in the second and third pandemic waves (6· 2 years, IQR 3· 1– 10· 1) versus first (3· 8 years, IQR 1· 7–8· 2; Supplementary Table 1). Most children were previously well with 16· 7% having a chronic comorbid condition including asthma (5· 9%), neurologic conditions (3· 0%), and obesity (3· 0%) (Supplementary Table 2). Comparisons between children meeting versus not meeting PIMS study criteria are presented in Supplementary Tables 3 and 4.

### Clinical, Laboratory and imaging features

Among all PIMS patients, the median total duration of fever was six days (IQR 5–8; Table 2). In addition to fever, 76· 8% of patients had gastrointestinal symptoms (i.e. abdominal pain, diarrhea, and/or vomiting), and more than half had mucocutaneous changes including rash (70· 9%), bilateral non-exudative conjunctivitis (70· 4%), and changes to the lips/oral cavity (64· 5%). Shock/hypotension was reported among 39· 2% of patients. Cases with positive SARS-CoV-2 linkage were more likely to experience gastrointestinal involvement than cases with a negative linkage (88· 6% vs. 63· 2%; p<0· 001), as well as shock/hypotension (60· 9% vs. 16· 0%; p<0· 001). Cases with negative SARS-CoV-2 linkage were more likely to have KD features including changes in lips/oral cavity (75· 5% vs. 56· 9%; p=0· 001) and peripheral extremities than cases with a positive SARS-CoV-2 linkage (61· 3% vs. 45· 0%, p=0· 007).

**Table 2.** Clinical, laboratory, and radiologic features of PIMS hospitalizations.

| Characteristics | All cases | SARS-CoV-2 linkage<sup>1</sup> |  |  |  |
| --- | --- | --- | --- | --- | --- |
|  |  | Positive | Negative | P value | Unknown |
| **Number of PIMS hospitalizations, N** | 406 | 202 | 106 | --- | 98 |
| **Duration of fever (days), median (IQR)<sup>2</sup>** | 6 (5–8) | 6 (5–8) | 7 (5–10) | 0.001 | 7 (5–8) |
| **Clinical features, n (%)** |  |  |  |  |  |
| Abdominal pain/vomiting/diarrhea | 312 (76.8) | 179 (88.6) | 67 (63.2) | <0.001 | 66 (67.3) |
| Rash | 288 (70.9) | 138 (68.3) | 74 (69.8) | 0.79 | 76 (77.6) |
| Bilateral bulbar conjunctival injection without exudate | 286 (70.4) | 148 (73.3) | 66 (62.3) | 0.046 | 72 (73.5) |
| Changes in lips/oral cavity | 262 (64.5) | 115 (56.9) | 80 (75.5) | 0.001 | 67 (68.4) |
| Changes in peripheral extremities | 219 (53.9) | 91 (45.0) | 65 (61.3) | 0.007 | 63 (64.3) |
| Shock/hypotension | 159 (39.2) | 123 (60.9) | 17 (16.0) | <0.001 | 19 (19.4) |
| Cervical lymphadenopathy >1.5 cm diameter | 103 (25.4) | 37 (18.3) | 32 (30.2) | 0.02 | 34 (34.7) |
| Periungual desquamation | 63 (15.5) | 16 (7.9) | 29 (27.4) | <0.001 | 18 (18.4) |
| **Coagulation dysfunction, n (%)** | 208 (51.2) | 134 (66.3) | 44 (41.5) | <0.001 | 30 (30.6) |
| **Laboratory features, median (IQR)** |  |  |  |  |  |
| Albumin (nadir; µmol/L) | 27 (23–33) | 24 (20–29) | 31 (26–36) | <0.001 | 29 (25–35) |
| ALT (peak; U/L) | 38 (19–90) | 46 (22–91) | 26 (16–85) | 0.41 | 38 (19–88) |
| AST (peak; U/L) | 49 (35–89) | 49 (36–95) | 46 (32–82) | 0.43 | 52 (37–102) |
| Bilirubin (peak; µmol/L) | 7.1 (4.9–14.0) | 9.2 (6.0–18.1) | 5.1 (3.5–10.0) | 0.09 | 6.0 (4.0–9.1) |
| CRP (peak; mg/L) | 147 (97–221) | 182 (118–239) | 118 (78–210) | <0.001 | 114 (79–180) |
| D-dimer (peak; µg/mL) | 2.1 (1.2–4.0) | 2.4 (1.5–4.3) | 1.9 (0.9–4.0) | 0.14 | 1.8 (1.1–3.5) |
| ESR (peak; mm/h) | 55 (41–78) | 50 (38–70) | 56 (45–75) | 0.28 | 60 (48–92) |
| Ferritin (peak; µg/L) | 322 (169–673) | 480 (276–1003) | 214 (117–444) | <0.001 | 175 (119–396) |
| LDH (peak; U/L) | 464 (317–796) | 402 (298–722) | 560 (303–832) | 0.14 | 535 (391–893) |
| Platelet count (nadir during admission; x10<sup>9</sup>/L) | 208 (134–321) | 147 (111–212) | 296 (207–364) | <0.001 | 288 (196–375) |
| Platelet count (peak after admission; x10<sup>9</sup>/L) | 496 (373–679) | 448 (329–609) | 561 (408–722) | 0.04 | 513 (406–688) |
| Sodium (nadir; µmol/L) | 134 (132–136) | 133 (130–136) | 135 (134–137) | <0.001 | 135 (133–137) |
| Troponin (peak; ng/L) | 13 (9–90) | 55 (10–219) | 10 (6–12) | <0.001 | 10 (5–12) |
| **Elevated laboratory features<sup>2</sup>, n / patients with tests (%)** |  |  |  |  |  |
| ALT > Institutional 'normal' reference | 210/369 (56.9) | 107/178 (60.1) | 51/103 (49.5) | 0.08 | 52/88 (59.1) |
| AST > Institutional 'normal' reference | 130/270 (48.1) | 67/135 (49.6) | 32/72 (44.4) | 0.48 | 31/63 (49.2) |
| Bilirubin > Institutional 'normal' reference | 37/174 (21.3) | 24/99 (24.2) | 9/38 (23.7) | 0.95 | <5/37 (<13.5) |
| CRP ≥ 30 mg/L | 396/405 (97.8) | 200/201 (99.5) | 101/106 (95.3) | 0.02 | 95/98 (96.9) |
| D-dimer > Institutional 'normal' reference | 317/331 (95.8) | 176/181 (97.2) | 77/83 (92.8) | 0.11 | 64/67 (95.5) |
| ESR ≥ 40 mm/h | 236/307 (76.9) | 99/140 (70.7) | 75/92 (81.5) | 0.06 | 62/75 (82.7) |
| Ferritin ≥ 500 μg/L | 132/381 (34.6) | 96/200 (48.0) | 21/101 (20.8) | <0.001 | 15/80 (18.8) |
| LDH > Institutional 'normal' reference | 162/309 (52.4) | 82/160 (51.2) | 52/89 (58.4) | 0.28 | 28/60 (46.7) |
| Troponin > Institutional 'normal' reference | 151/340 (44.4) | 123/192 (64.1) | 18/88 (20.5) | <0.001 | 10/60 (16.7) |
| **Abnormal echocardiogram, n / patients with echo (%)<sup>3</sup>** | 191/392 (48.7) | 117/199 (58.8) | 37/99 (37.4) | <0.001 | 37/94 (39.4) |
| Decreased heart function | 71/392 (18.1) | 61–64/199 (30.7–32.2) | 5–8/99 (5.1–8.1) | <0.001 | <5/94 (<5.3) |
| Coronary artery lesions, Z-score ≥ 2.5 | 66/392 (16.8) | 28/199 (14.1) | 21/99 (21.2) | 0.12 | 17/94 (18.1) |
| Coronary artery lesions, Z-score < 2.5 | 56/392 (14.3) | 27/199 (13.6) | 11/99 (11.1) | 0.55 | 18/94 (19.1) |
| Pericardial effusion | 43/392 (11.0) | 30/199 (15.1) | 7/99 (7.1) | 0.048 | 6/94 (6.4) |
| Myocarditis | 40/392 (10.2) | 35/199 (17.6) | <5/99 (<5.1) | <0.001 | <5/94 (<5.3) |
| Valvular insufficiency | 33/392 (8.4) | 23–26/199 (11.6–13.1) | <5/99 (<5.1) | 0.008 | 5–8/94 (5.3–8.5) |
| **Radiologic findings, n / patients with imaging (%)** |  |  |  |  |  |
| Abnormal chest x-ray | 93/282 (33.0) | 66/166 (39.8) | 16/72 (22.2) | 0.009 | 11/44 (25.0) |
| Abnormal CT scan | 24/44 (54.5) | DNS | DNS | --- | DNS |
| Abnormal MRI scan | 8/23 (34.8) | DNS | DNS | --- | DNS |
| Abnormal ultrasound | 48/114 (42.1) | 28/57 (49.1) | 12/30 (40.0) | 0.42 | 8/27 (29.6) |

ALT = Alanine aminotransferase test; AST = Aspartate aminotransferase test; CRP = C-reactive protein; CT = Computed tomography; ESR = Erythrocyte sedimentation rate; LDH = Lactate dehydrogenase; MRI = Magnetic resonance imaging.
<sup>1</sup>A positive link indicates a positive PCR or rapid test, serology, or close contact; a negative link indicates a negative serology result with no other positive test or close contact; an unknown link indicates a lack of a positive test or close contact, with serology either not conducted or not reported.
<sup>2</sup>Data available for 386 cases (188 with positive link, 104 with negative link, and 94 with unknown link).
<sup>3</sup>Patients may have multiple cardiac findings, therefore categories exceed total.

Among all PIMS cases, the median CRP peak was 147 mg/L (IQR 97–221) and median ESR peak was 55 mm/h (IQR 41–78; Table 2). Cases with positive SARS-CoV-2 linkages had significantly higher CRP and ferritin, while having significantly lower sodium, platelet, and albumin nadirs, compared to cases with negative linkage (all p-values <0· 001). One third of PIMS cases had abnormal chest radiographs, with common findings including pulmonary opacities, pulmonary edema, peribronchial thickening and pleural effusion.

Among PIMS patients for whom echocardiograms were conducted (n=392), 48· 7% had cardiac involvement (Table 2). Most common were coronary artery abnormalities (26· 5%), among whom the median maximum Z-score was 3· 0 (IQR 2· 4–3· 7) at the time of report. Decreased heart function was also commonly reported (18· 1%), with a median minimum ejection fraction of 45% (IQR 40–50%). Children <1 year old more commonly had coronary artery abnormalities with Z-scores ≥ 2· 5 than older age groups (38· 1% vs. 13· 2–15· 7%; p=0· 001), while children aged 6–17 more commonly had decreased heart function than younger age groups (31· 3–32· 1% vs. 0· 0–7· 4%; p<0· 001) (Supplementary Table 5). In those who had serum troponin levels measured, it was elevated in 44· 4% with a median peak of 13 ng/L (IQR 9–90). Compared to cases with negative SARS-CoV-2 linkage, cases with positive SARS-CoV-2 linkage more commonly experienced decreased heart function (30· 7–32· 2% versus 5· 1-8· 1%; p<0· 001) and elevated troponin (64· 1% versus 20· 5%; p<0· 001), but no difference in coronary artery lesions (Table 2).

### Treatments and supports

Overall, 28· 6% of children required some form of respiratory or hemodynamic support, including more frequently in cases with positive SARS-CoV-2 linkage than those with negative linkage (49· 0% vs. 9· 4%; p<0· 001; Table 3). Respiratory or hemodynamic supports were also more frequently required during second/third waves (34· 4% vs. 14· 0% in wave one; p<0· 001) (Supplementary Table 1). Supports included vasopressors (20· 0% of patients), low-flow oxygen (13· 1%), high-flow nasal cannula (5· 2%), conventional mechanical (3· 4%), and non-invasive ventilation (3· 2%). No patients received extracorporeal membrane oxygenation. Most PIMS patients (94· 3%) received intravenous immunoglobulin G (IVIG; with 80· 4% receiving one dose and 13· 9% receiving two or more doses), and 67· 0% received systemic corticosteroids. Together, 65· 8% received both IVIG and steroids, 28· 6% received IVIG only, 1· 2% received corticosteroids only, and 4· 4% received neither IVIG nor steroids, while 5· 9% of patients received a biologic agent. Most patients received aspirin (90· 1%) and 11· 3% received either prophylactic or therapeutic anticoagulation. Forty-five patients (11· 1%) had a concurrent infection determined to have contributed to clinical findings, though PIMS remained the primary cause of hospitalization (Supplementary Table 6).

**Table 3.** Supports and treatments administered to patients hospitalized with PIMS.

| Characteristics, n (%) | All cases | SARS-CoV-2 linkage<sup>1</sup> |  |  |  |
| --- | --- | --- | --- | --- | --- |
|  |  | Positive | Negative | P value | Unknown |
| **Number of PIMS hospitalizations, N** | 406 | 202 | 106 | --- | 98 |
| **Respiratory/hemodynamic support required, n (%)<sup>2</sup>** | 116 (28.6) | 99 (49.0) | 10 (9.4) | <0.001 | 7 (7.1) |
| Vasopressors | 81 (20.0) | 74 (36.6) | <5 (<4.7) | <0.001 | <5 (<5.1) |
| Low-flow oxygen | 53 (13.1) | 46 (22.8) | <5 (<4.7) | <0.001 | <5 (<5.1) |
| High-flow nasal cannula | 21 (5.2) | 17–20 (8.4–9.9) | <5 (<4.7) | 0.01 | 0 (0.0) |
| Conventional mechanical ventilation | 14 (3.4) | 9–12 (4.5–5.9) | <5 (<4.7) | 0.04 | <5 (<5.1) |
| Non-invasive ventilation (e.g. CPAP/BiPAP) | 13 (3.2) | 9–12 (4.5–5.9) | <5 (<4.7) | 0.55 | 0 (0.0) |
| **Treatments received** |  |  |  |  |  |
| Immunoglobulin (IVIG)<sup>3</sup> | 383 (94.3) | 197 (97.5) | 99 (93.4) | 0.12 | 87 (88.8) |
| Aspirin | 366 (90.1) | 182 (90.1) | 98 (92.5) | 0.49 | 86 (87.8) |
| Steroids<sup>3</sup> | 272 (67.0) | 167 (82.7) | 58 (54.7) | <0.001 | 47 (48.0) |
| Antibiotics | 236 (58.1) | 146 (72.3) | 48 (45.3) | <0.001 | 42 (42.9) |
| Anticoagulation<sup>4</sup> | 46 (11.3) | 34–37 (16.8–18.3) | 8–11 (7.5–10.4) | 0.02 | <5 (<5.1) |
| Biologics<sup>4</sup> | 24 (5.9) | 17 (8.4) | <5 (<4.7) | 0.06 | <5 (<5.1) |
| Antivirals<sup>4</sup> | <5 (<1.2) | DNS | DNS | --- | DNS |
| **IVIG doses received<sup>5</sup>** |  |  |  | 0.07 |  |
| Zero | 23 (5.7) | 5 (2.5) | 7 (6.6) |  | 11 (11.6) |
| One | 324 (80.4) | 177 (87.6) | 83 (78.3) |  | 64 (67.4) |
| ≥Two | 56 (13.9) | 20 (9.9) | 16 (15.1) |  | 20 (21.1) |

DNS = Data not shown due to <5 cell counts in one or both subgroups.
<sup>1</sup>A positive link indicates a positive PCR or rapid test, serology, or close contact; a negative link indicates a negative serology result with no other positive test or close contact; an unknown link indicates a lack of a positive test or close contact, with serology either not conducted or not reported.
<sup>2</sup>Patients may have received multiple supports, therefore sum of categories exceed total.
<sup>3</sup>Includes 267 receiving both IVIG and steroids, 116 receiving IVIG only, 5 receiving steroids only, and 18 receiving neither IVIG nor steroids.
<sup>4</sup>Anticoagulation includes prophylactic and therapeutic anticoagulation. Biologics includes anti-TNF, anti-IL-1, and anti-IL-6.
<sup>5</sup>Number of IVIG doses not available for three participants (3 with unknown link).

There were 127 (31· 3%) PIMS patients admitted to the ICU, among whom the median ICU length of stay was three days (IQR 2–4). No deaths due to PIMS occurred (Table 1). Children with positive SARS-CoV-2 linkage were 2· 7 times (95% CI 1· 6–4· 5) more likely to be admitted to ICU than children with negative linkage (Table 4). Compared to children aged 1–5 years, older age was also associated with greater risk of ICU admission for ages 6–12 years (aRR 2· 5 [1· 7–3· 6]) and ages 13–17 years (aRR 2· 3 [1· 5–3· 5]). We found no significant association with age under one year, sex, or co-morbidities, and higher risk among the second and third waves in crude analyses was attenuated in multivariable analysis. When comparing clinical features among children by level of care required (i.e. ICU vs. inpatient ward; Supplementary Table 7), children admitted to ICU were more likely to experience gastrointestinal involvement (92· 2% vs. 70· 5%; p<0· 001), and shock/hypotension (83· 5% vs. 19%; p<0· 001).

**Table 4.**
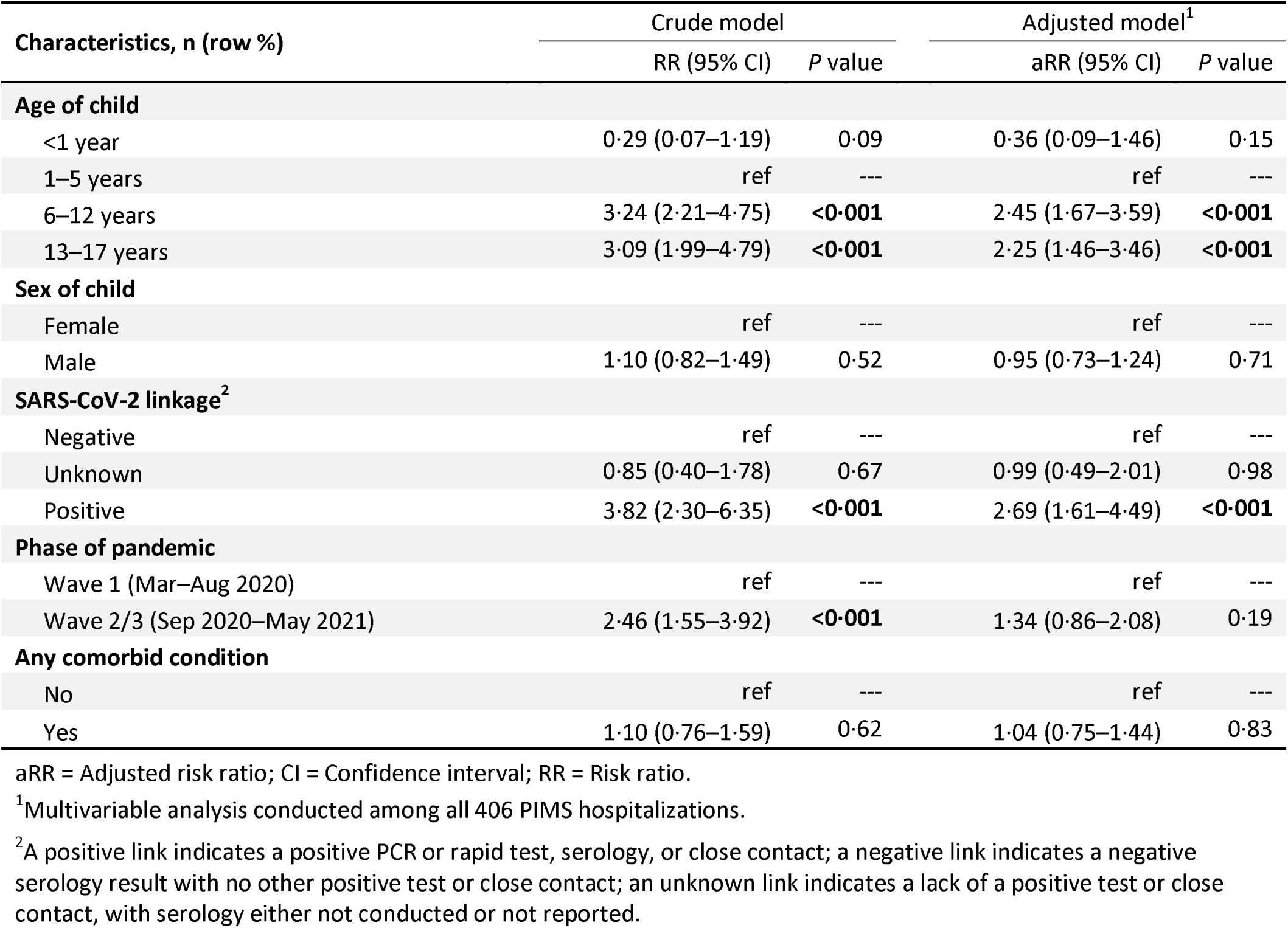
Multivariable Poisson regression analysis of predictors of ICU admission.

### Time series and Epidemiology

Accounting for <18-year population denominators, the CPSP reported cases corresponds to a minimum study period incidence of 5· 6 PIMS hospitalizations (95% CI 5· 1–6· 2) and 2· 8 PIMS hospitalizations with a positive SARS-CoV-2 link (95% CI 2· 4–3· 2) per 100,000 population (Supplementary Table 8). PIMS hospitalizations peaked nationally first in May 2020 and again in January 2021, and were most strongly correlated with a five-week lag behind Canadian SARS-CoV-2 case counts (Spearman’s rho 0· 68) (Figure 2). Most PIMS hospitalizations occurred in Ontario (39· 9%), Quebec (36· 5%), and Alberta (12· 6%). A higher proportion of cases hospitalized in the second/third waves had SARS-CoV-2 linkages (61· 6%) compared to the first wave (18· 8%, p<0· 001; Supplementary Table 1).

**Figure 2.**
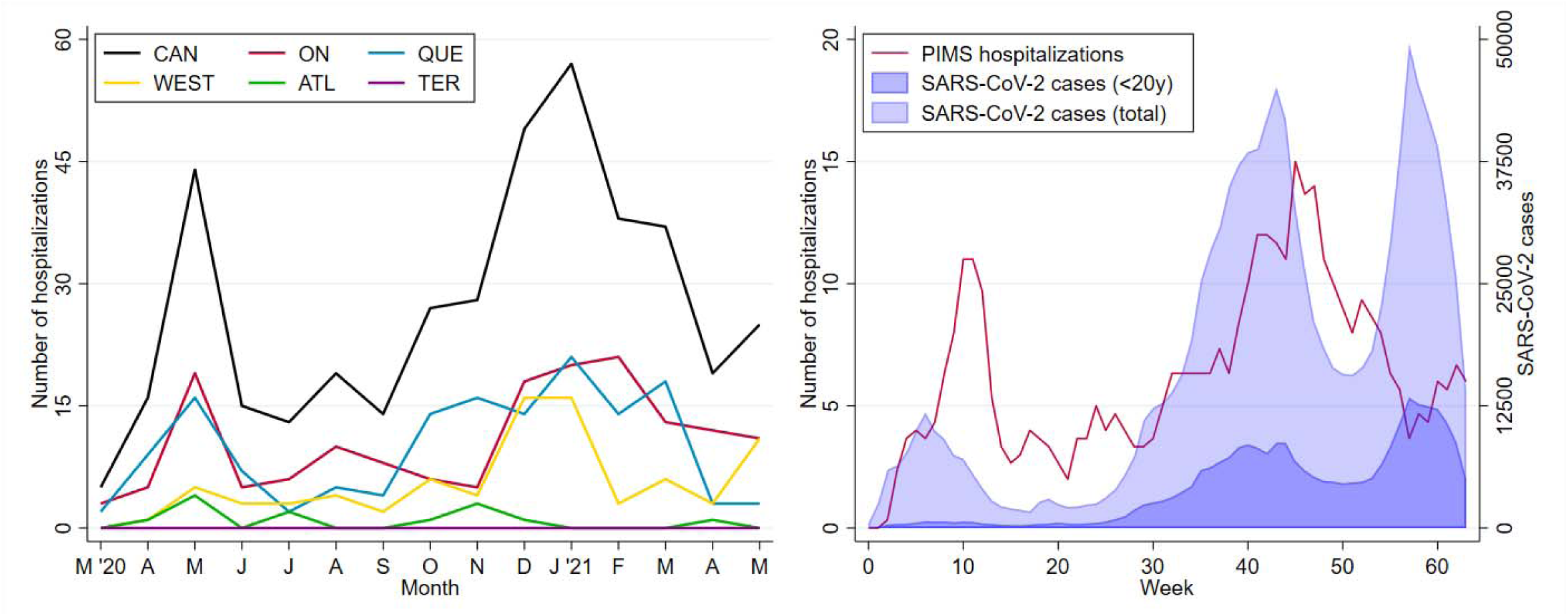
Time series of a) monthly PIMS admissions by region (left); and b) weekly PIMS admissions and all <20-year Canadian cases of COVID-19 (right). Footnotes: Panel A includes 406 hospitalizations from across Canada (CAN), including 162 from Ontario (ON), 148 from Quebec (QUE), 83 from Western Canada (WEST; 51 from Alberta, 15 from British Columbia, and 17 from Manitoba or Saskatchewan); 13 from Atlantic Canada (ATL; including New Brunswick, Newfoundland and Labrador, Nova Scotia, or Prince Edward Island); and zero from Territories (TER; including Northwest Territories, Nunavut, Yukon). In panel B, PIMS data represent the three-week moving average of weekly hospitalizations. Week zero is defined as the week beginning March 2, 2020 and January 1, 2021 occurs in week 43. COVID-19 cases were extracted from https://health-infobase.canada.ca/covid-19/epidemiological-summary-covid-19-cases.html, and reflect the date of illness onset.

## Discussion

Leveraging established national public health surveillance infrastructure using physician reported data, this study describes 406 hospitalized children with PIMS across Canada from May 2020 to May 2021. By comparing PIMS cases with positive and negative SARS-CoV-2 linkages, we were able to study the impact of SARS-CoV-2 linkage on clinical and laboratory features, and outcomes in hospitalized children presenting with presumed post-infectious hyperinflammatory syndrome during the first three waves of the COVID-19 pandemic. It is important to note that as waves of SARS-CoV-2 continue to infect increasingly large proportions of populations around the world and paediatric vaccination increases, the utility of current serologic tests in the diagnosis of post-SARS-CoV-2 conditions and clinical decision-making will lessen.

Compared to children with negative SARS-CoV-2 linkage, children with positive SARS-CoV-2 linkage were older, had more severe gastrointestinal and cardiac involvement (decreased heart function, shock), a more hyperinflammatory laboratory picture (higher CRP, ferritin, and troponin peak levels with coagulation dysfunction), higher rates of ICU admission, requirements of respiratory/hemodynamic support, and were more likely to receive corticosteroids and antibiotics. Our study findings for this group are in line with descriptive case series describing clinical signs and symptoms of PIMS and MIS-C regardless of case definition, yet most comparisons are made with COVID-19 patients and/or pre-pandemic KD.^15,16^ The number of PIMS hospitalizations and ICU admissions was also consistent with a Canadian national case series describing high-level MIS-C outcomes.^17^

Conversely, children with negative SARS-CoV-2 linkage were younger, experienced more KD features including changes in lips/oral cavity and peripheral extremities, and a higher platelet peak after admission. While the younger age and thrombocytosis seen in this group may be consistent with a KD phenotype, there are several features that suggest at least some of these cases are not entirely typical of pre-pandemic KD. The incidence of shock (16%) in this group is more than twice that reported in KD shock syndrome (7%).^12^ Secondly, approximately 10-20% of patients with KD have recalcitrant fever after their first IVIG infusion, also known as being “IVIG resistant”.^12^ In our study, more than half of these cases (54· 7%) were treated with corticosteroids, with 15· 1% receiving ≥ 2 IVIG infusions. While the order in which immunomodulatory agents were given (i.e. concurrently or sequentially) was not captured in this study, these combined treatments surpass the rates of recalcitrant KD.^12^ Of note, the American College of Rheumatology task force first recommended that IVIG and glucocorticoids could be used alone or in combination for treatment of this syndrome in July 2020 (Version 1), and by April 2021 (Version 2), recommended upfront steroids. This may partly explain higher steroid use in those patients.^18,19^ Therefore, although this group did not have evidence of SARS-CoV-2 linkage, many still experienced the severe end of the disease spectrum. While SARS-CoV-2 linkage is an important epidemiologic feature, requiring it in a case definition may be overly specific and result in missing certain cases. A recent study showed that some individuals do not develop a serologic response because of pre-existing memory T-cell responses, with cross-protective potential against SARS-CoV-2.^20^ Additional study is needed to better understand the post-infectious hyperinflammatory syndrome spectrum and the heterogeneous range of phenotypes described and relationship with the infectious trigger.

Thirty percent of PIMS cases required ICU admission, emphasizing the importance of identifying potential risk factors for ICU admission that might inform prognosis and early intervention. Our study showed an association with older age groups (6-12 and 13-17 years) and SARS-CoV-2 linkage but no association with sex or underlying comorbidities. Furthermore, the identified risk factors above are consistent with trends seen in previous literature.^21^ It remains unclear why there may be a possible predilection towards older children, though some theories related to age-based immune response and susceptibility have been proposed.^5,15,19,22,23^ Though confounded by the PIMS-associated age distribution, the reported numbers of specific comorbid conditions in our study including asthma (5· 9%), obesity (3· 0%), and neurologic/neurodevelopmental conditions (3· 0%) fall within reported prevalence in the general paediatric population.^24,25^ Five children (1· 2%) had prior history of KD which is in keeping with the reported percentage of recurrence of KD pre-pandemic that could vary among different ethnicities.^12^

During the study period, the minimum national incidence of PIMS hospitalization was 5· 6 per 100,000 population and for ICU admission was 1· 8 per 100,000 population (2· 8 and 1· 4 per 100,000 population for cases with positive SARS-CoV-2 linkages, respectively) which are comparable to that of acute COVID-19 disease ascertained in the same CPSP study (4· 6 hospitalizations and 0· 8 ICU admissions per 100,000 population, excluding incidental SARS-CoV-2).^10^ The incidence of PIMS hospitalizations with SARS-CoV-2 linkages was highest in those aged 1–5 years (3· 3 per 100,000 population) and those aged 6–12 years (3· 1 per 100,000 population). The estimate of worldwide incidence of this syndrome is still unknown. Although PIMS is a rare complication of SARS-CoV-2 infection, these figures underscore an important consequence of the COVID-19 pandemic in the paediatric population. Reassuringly, observational studies have shown overall low mortality in PIMS with relatively rapid recovery of organ dysfunction with appropriate timely treatment, though longitudinal follow-up is limited.^26,27^ It also supports the importance of vaccination of eligible children as early evidence supports this to be effective at preventing PIMS in adolescents.^28^

While our study cannot establish causality, the findings showed that peaks of PIMS hospitalizations correlated with a temporal lag of five weeks after peaks in all Canadian SARS-CoV-2 infections, similarly reported in other studies ranging from 2-6 weeks.^2,15^ Higher peaks were seen in three Canadian jurisdictions (Ontario, Quebec, and Alberta) with the highest numbers of acute SARS-CoV-2 infections. This supports a possible post-infectious immune dysregulation phenomenon.^29^ The higher numbers of cases witnessed in second/third waves vs. first (Supplementary table 1) may be partly reflect the real-time learning curve of health care providers in better identifying cases. This also highlights the success of this voluntary reporting system.

This study has a few limitations. The voluntary nature of CPSP reporting means that not all cases may have been reported. Second, the online PIMS case report form was developed soon after the first identification of the clinical entity, therefore data on other important clinical or laboratory markers such as NT-proBNP and lymphopenia were not included in the study. In addition, several indicators ascertained by physician report, including population group of the child and cardiac findings such as myocarditis and shock were not based on pre-defined diagnostic criteria.

## Conclusion

While PIMS is rare, it remains an important consequence of SARS-CoV-2 infection in children in which almost 1 in 3 hospitalized children required ICU admission and respiratory/hemodynamic support. At– risk groups for ICU admission include children 6 years and older and those with SARS-CoV-2 linkages. These results provide baseline data prior to implementation of SARS-CoV-2 vaccination for children and can be used to monitor changes to the epidemiology of PIMS as vaccination in different age groups is introduced. We demonstrate that in the milieu of widespread SARS-CoV-2 transmission, post-infectious hyperinflammatory phenotypes observed are more severe regardless of recognized SARS-CoV-2 exposure compared to pre-pandemic KD. Collectively, these findings help both better define the epidemiology of this condition in Canada and serve to inform clinical practice and public health responses to PIMS.

## Data Availability

De-identified data that underlie the results reported in this article (text, tables, figures and appendices) and that abide by the privacy rules of the Canadian Paediatric Surveillance Program and the Public Health Agency of Canada can be made available to investigators whose secondary data analysis study protocol has been approved by an independent research ethics board.

## Author contributions

The study was conceived by TET, M-PM, SKM, DSF, EH, RS, and RSMY. TET write the first draft of the manuscript and DSF conducted the statistical analysis. All authors had access to the data, and DSF, CMH, ML, MK, and SKM have accessed and verified the data underlying the study. All authors contributed to data collection, reviewed the study results and manuscript, and approved the final manuscript.

## Acknowledgements

The authors wish to thank the paediatricians, paediatric sub-specialists, and health professionals who voluntarily responded to CPSP surveys. We also wish to thank the members and leadership of the Paediatric Inpatient Research Network (PIRN) for cases reported and their dedication to the CPSP. We are enormously grateful to the staff of the CPSP for their dedication, diligence, and commitment to this study. Lastly, the authors also wish to thank members of the CPSP Scientific Steering Committee who serve as stewards of the program.

## Supplementary Tables

**Supplementary Table 1.** Characteristics of PIMS hospitalizations by pandemic wave.

| Characteristics | Phase of Pandemic |  | P value |
| --- | --- | --- | --- |
|  | First Wave<br>(Mar–Aug 2020) | Second/Third Wave<br>(Sep 2020–May 2021) |  |
| **Number of PIMS hospitalizations, N** | 112 | 294 | --- |
| **Age (years), median (IQR)** | 3·8 (1·7–8·2) | 6·2 (3·1–10·1) | 0·001 |
| **SARS-CoV-2 linkage, n (%)<sup>1</sup>** |  |  | <0·001 |
| Positive | 21 (18·8) | 181 (61·6) | --- |
| Negative | 38 (33·9) | 68 (23·1) | --- |
| Unknown | 53 (47·3) | 45 (15·3) | --- |
| **Positive SARS-CoV-2 linkage, n (%)** |  |  |  |
| Close contact with confirmed SARS-CoV-2 | 13 (11·6) | 122 (41·5) | <0·001 |
| PCR positive | 7 (6·3) | 98 (33·3) | <0·001 |
| Serology positive | 5 (4·5) | 92 (31·3) | <0·001 |
| **History of negative serology, n(%)** | 39 (34·8) | 77 (26·2) | 0·09 |
| **Treatments received** |  |  |  |
| Immunoglobulin (IVIG) | 99 (88·4) | 284 (96·6) | 0·001 |
| Aspirin | 96 (85·7) | 270 (91·8) | 0·06 |
| Steroids | 62 (55·4) | 210 (71·4) | 0·002 |
| Antibiotics | 55 (49·1) | 181 (61·6) | 0·02 |
| Anticoagulation<sup>2</sup> | 6 (5·4) | 40 (13·6) | 0·02 |
| Biologics<sup>2</sup> | 5 (4·5) | 19 (6·5) | 0·45 |
| **Outcomes** |  |  |  |
| Abnormal echocardiogram, n / patients with echo (%) | 41/110 (37·3) | 150/282 (53·2) | 0·005 |
| Respiratory/hemodynamic support required, n (%) | 15 (13·4) | 101 (34·4) | <0·001 |
| Admitted to ICU, n (%) | 17 (15·2) | 110 (37·4) | <0·001 |
| Death, n (%) | 0 (0·0) | 0 (0·0) | --- |

ICU = Intensive care unit; IQR = Interquartile range; IVIG = Intravenous immunoglobulin; PCR = Polymerase chain reaction
<sup>1</sup>A positive link indicates a positive PCR or rapid test, serology, or close contact; a negative link indicates a negative serology result with no other positive test or close contact; an unknown link indicates a lack of a positive test or close contact, with serology either not conducted or not reported.
<sup>2</sup>Anticoagulation includes prophylactic and therapeutic anticoagulation. Biologics includes anti-TNF, anti-IL-1, and anti-IL-6.

**Supplementary Table 2.** Chronic comorbidities among patients with PIMS.

| Characteristics, n (%) | Frequency (N) | Percent (%) |
| --- | --- | --- |
| **Number of PIMS hospitalizations, N** | 406 | --- |
| **Prior history of Kawasaki disease** | 5 | 1.2 |
| **Born prematurely (<37 weeks')<sup>2</sup>** | 11 | 2.9 |
| **Any comorbid conditions** |  |  |
| None/Unknown | 338 | 83.3 |
| One or more | 68 | 16.7 |
| **Specific comorbid conditions** |  |  |
| Asthma | 24 | 5.9 |
| Neurologic/neurodevelopmental condition | 12 | 3.0 |
| Obesity | 12 | 3.0 |
| Sickle cell/other hematologic condition | 9 | 2.2 |
| Congenital heart disease | 5 | 1.2 |
| Genetic or metabolic disease | 5 | 1.2 |
| Chronic rheumatologic disease | <5 | <1.2 |
| Chronic lung disease (excluding asthma) | <5 | <1.2 |
| Chronic renal disease | <5 | <1.2 |
| Diabetes | <5 | <1.2 |
| Gastrointestinal or liver disease | <5 | <1.2 |
| Immunosuppression | <5 | <1.2 |
| Malignancy | 0 | 0.0 |
| Transplant recipient | 0 | 0.0 |

<sup>2</sup>Prematurity status known for 395 patients.

**Supplementary Table 3.** Demographic and outcome characteristics of children who met versus did not meet the study PIMS case definition.

| Characteristics, n (%) | Met PIMS criteria |  | *P* value |
| --- | --- | --- | --- |
|  | Yes | No |  |
| **Number of patients, N** | 406 | 81 | --- |
| **Age of child (years)<sup>1</sup>** |  |  | 0.006 |
| <1 year | 43 (10.6) | 18 (23.7) | --- |
| 1–5 years | 170 (41.9) | 34 (44.7) | --- |
| 6–12 years | 138 (34.0) | 16 (21.1) | --- |
| 13–17 years | 55 (13.5) | 8 (10.5) | --- |
| **Sex of child** |  |  | 0.36 |
| Female | 163 (40.1) | 37 (45.7) | --- |
| Male | 243 (59.9) | 44 (54.3) | --- |
| **Positive SARS-CoV-2 linkage<sup>2</sup>** | 202 (49.8) | 25 (30.9) | 0.002 |
| **Phase of pandemic** |  |  | 0.001 |
| Wave 1 (Mar–Aug 2020) | 112 (27.6) | 37 (45.7) | --- |
| Wave 2 (Sep 2020–Mar 2021) | 294 (72.4) | 44 (54.3) | --- |
| **Admitted to ICU** | 127 (31.3) | 12 (14.8) | 0.003 |

<sup>1</sup>Age unavailable for five children who did not meet PIMS criteria.
<sup>2</sup>Defined as a positive PCR or rapid test, serology, or close contact.

**Supplementary Table 4.** Clinical, laboratory, and radiologic features of children who met versus did not meet the study PIMS case definition.

| Characteristics | Met PIMS criteria |  | P value |
| --- | --- | --- | --- |
|  | Yes | No |  |
| **Number of patients, N** | 406 | 81 | --- |
| **Duration of fever (days), median (IQR)<sup>1</sup>** | 6 (5–8) | 6 (5–9) | 0.43 |
| **Clinical features, n (%)** |  |  |  |
| Abdominal pain/vomiting/diarrhea | 312 (76.8) | 43 (53.1) | <0.001 |
| Rash | 288 (70.9) | 27 (33.3) | <0.001 |
| Bilateral bulbar conjunctival injection without exudate | 286 (70.4) | 14 (17.3) | <0.001 |
| Changes in lips/oral cavity | 262 (64.5) | 19 (23.5) | <0.001 |
| Changes in peripheral extremities | 219 (53.9) | 20 (24.7) | <0.001 |
| Shock/hypotension | 159 (39.2) | 11 (13.6) | <0.001 |
| Cervical lymphadenopathy >1.5 cm diameter | 103 (25.4) | 5 (6.2) | <0.001 |
| Periungual desquamation | 63 (15.5) | <5 (<6.2) | 0.00 |
| **Coagulation dysfunction, n (%)** | 208 (51.2) | 29 (35.8) | 0.011 |
| **Laboratory features, median (IQR)** |  |  |  |
| Albumin (nadir; µmol/L) | 27 (23–33) | 32 (27–38) | 0.001 |
| ALT (peak; U/L) | 38 (19–90) | 21 (15–54) | 0.008 |
| AST (peak; U/L) | 49 (35–89) | 43 (32–58) | 0.20 |
| Bilirubin (peak; µmol/L) | 7.1 (4.9–14.0) | 8.1 (3.0–16.0) | 0.78 |
| CRP (peak; mg/L) | 147 (97–221) | 129 (53–201) | <0.001 |
| D-dimer (peak; µg/mL) | 2.1 (1.2–4.0) | 1.6 (1.0–3.2) | 0.27 |
| ESR (peak; mm/h) | 55 (41–78) | 55 (44–81) | 0.31 |
| Ferritin (peak; µg/L) | 322 (169–673) | 282 (173–690) | 0.91 |
| LDH (peak; U/L) | 464 (317–796) | 725 (450–1013) | 0.001 |
| Platelet count (nadir during admission; x10<sup>9</sup>/L) | 208 (134–321) | 212 (158–325) | 0.47 |
| Platelet count (peak after admission; x10<sup>9</sup>/L) | 496 (373–679) | 474 (360–575) | 0.08 |
| Sodium (nadir; µmol/L) | 134 (132–136) | 136 (134–138) | 0.006 |
| Troponin (peak; ng/L) | 13 (9–90) | 10 (10–10) | 0.03 |
| **Elevated laboratory features, n / patients with tests (%)** |  |  |  |
| ALT > Institutional 'normal' reference | 210/369 (56.9) | 27/65 (41.5) | 0.02 |
| AST > Institutional 'normal' reference | 130/270 (48.1) | 19/48 (39.6) | 0.27 |
| Bilirubin > Institutional 'normal' reference | 37/174 (21.3) | 5/22 (22.7) | 0.79 |
| CRP ≥ 30 mg/L | 396/405 (97.8) | 55/70 (78.6) | <0.001 |
| D-dimer > Institutional 'normal' reference | 317/331 (95.8) | 59/61 (96.7) | >0.99 |
| ESR ≥ 40 mm/h | 236/307 (76.9) | 46/61 (75.4) | 0.81 |
| Ferritin ≥ 500 µg/L | 132/381 (34.6) | 18/65 (27.7) | 0.27 |
| LDH > Institutional 'normal' reference | 162/309 (52.4) | 31/55 (56.4) | 0.59 |
| Troponin > Institutional 'normal' reference | 151/340 (44.4) | 7/57 (12.3) | <0.001 |
| **Abnormal echocardiogram, n / patients with echo (%)<sup>2</sup>** | 191/392 (48.7) | 14/70 (20.0) | <0.001 |
| Decreased heart function | 71/392 (18.1) | 5/70 (7.1) | 0.02 |
| Coronary artery lesions, Z-score ≥ 2.5 | 66/392 (16.8) | <5/70 (<7.1) | 0.02 |
| Coronary artery lesions, Z-score < 2.5 | 56/392 (14.3) | <5/70 (<7.1) | 0.02 |
| Pericardial effusion | 43/392 (11.0) | <5/70 (<7.1) | 0.03 |
| Myocarditis | 40/392 (10.2) | <5/70 (<7.1) | 0.049 |
| Valvular insufficiency | 33/392 (8.4) | <5/70 (<7.1) | 0.23 |
| **Radiologic findings, n / patients with tests (%)** |  |  |  |
| Abnormal chest x-ray | 93/282 (33.0) | 12/48 (25.0) | 0.27 |
| Abnormal ultrasound | 48/114 (42.1) | 7/18 (38.9) | 0.80 |

ALT = Alanine aminotransferase test; AST = Aspartate aminotransferase test; CRP = C-reactive protein; ESR = Erythrocyte sedimentation rate; LDH = Lactate dehydrogenase.
<sup>1</sup>Data available for 386 cases meeting criteria and 72 cases not meeting criteria.
<sup>2</sup>Patients may have multiple cardiac findings, therefore categories exceed total.

**Supplementary Table 5.** Clinical, laboratory, and radiologic features of PIMS hospitalizations by age group.

| Characteristics | Child age category |  |  |  | P value |
| --- | --- | --- | --- | --- | --- |
|  | <1 year | 1–5 years | 6–12 years | 13–17 years |  |
| **Number of PIMS hospitalizations, N** | 43 | 170 | 138 | 55 | --- |
| **Positive SARS-CoV-2 linkage, n (%)** | 9 (20.9) | 64 (37.6) | 90 (65.2) | 39 (70.9) | <0.001 |
| **History of negative serology, n (%)** | 16 (37.2) | 59 (34.7) | 31 (22.5) | 10 (18.2) | 0.02 |
| **Duration of fever (days), median (IQR)<sup>1</sup>** | 7 (5–8) | 7 (5–9) | 6 (5–8) | 6 (4–8) | 0.26 |
| **Clinical features, n (%)** |  |  |  |  |  |
| Rash | 35 (81.4) | 128 (75.3) | 90 (65.2) | 35 (63.6) | 0.06 |
| Bilateral bulbar conjunctival injection without exudate | 33 (76.7) | 125 (73.5) | 93 (67.4) | 35 (63.6) | 0.33 |
| Abdominal pain/vomiting/diarrhea | 25 (58.1) | 116 (68.2) | 123 (89.1) | 48 (87.3) | <0.001 |
| Changes in lips/oral cavity | 25 (58.1) | 128 (75.3) | 80 (58.0) | 29 (52.7) | 0.002 |
| Changes in peripheral extremities | 22 (51.2) | 103 (60.6) | 70 (50.7) | 24 (43.6) | 0.11 |
| Periungual desquamation | 6 (14.0) | 35 (20.6) | 15 (10.9) | 7 (12.7) | 0.11 |
| Cervical lymphadenopathy >1.5 cm diameter | 5 (11.6) | 61 (35.9) | 30 (21.7) | 7 (12.7) | <0.001 |
| Shock/hypotension | <5 (<11.6) | 37–40 (21.8–23.5) | 80–83 (58.0–60.1) | 34–37 (61.8–67.3) | <0.001 |
| **Coagulation dysfunction, n (%)** | 18 (41.9) | 61 (35.9) | 92 (66.7) | 37 (67.3) | <0.001 |
| **Abnormal echocardiogram, n / patients with echo (%)<sup>2</sup>** | 20/42 (47.6) | 58/163 (35.6) | 80/134 (59.7) | 33/53 (62.3) | <0.001 |
| Coronary artery lesions, Z-score $\geq 2.5$ | 16/42 (38.1) | 22/163 (13.5) | 21/134 (15.7) | 7/53 (13.2) | 0.001 |
| Coronary artery lesions, Z-score < 2.5 | 10/42 (23.8) | 21/163 (12.9) | 16/134 (11.9) | 9/53 (17.0) | 0.23 |
| Decreased heart function | 0/42 (0.0) | 12/163 (7.4) | 42/134 (31.3) | 17/53 (32.1) | <0.001 |
| Myocarditis | 0/42 (0.0) | <5/163 (<3.1) | 24–27/134 (17.9–20.1) | 12–15/53 (22.6–28.3) | <0.001 |
| Pericardial effusion | 0/42 (0.0) | 12/163 (7.4) | 22/134 (16.4) | 9/53 (17.0) | 0.002 |
| Valvular insufficiency | 0/42 (0.0) | 12–15/163 (7.4–9.2) | 17–20/134 (12.7–14.9) | <5/53 (<9.4) | 0.047 |

<sup>1</sup>Data available for 41 cases aged <1 year, 164 aged 1–5 years, 131 aged 6–12 years, and 50 aged 13–17 years.<sup>2</sup>Patients may have multiple cardiac findings, therefore categories exceed total.

**Supplementary Table 6.**
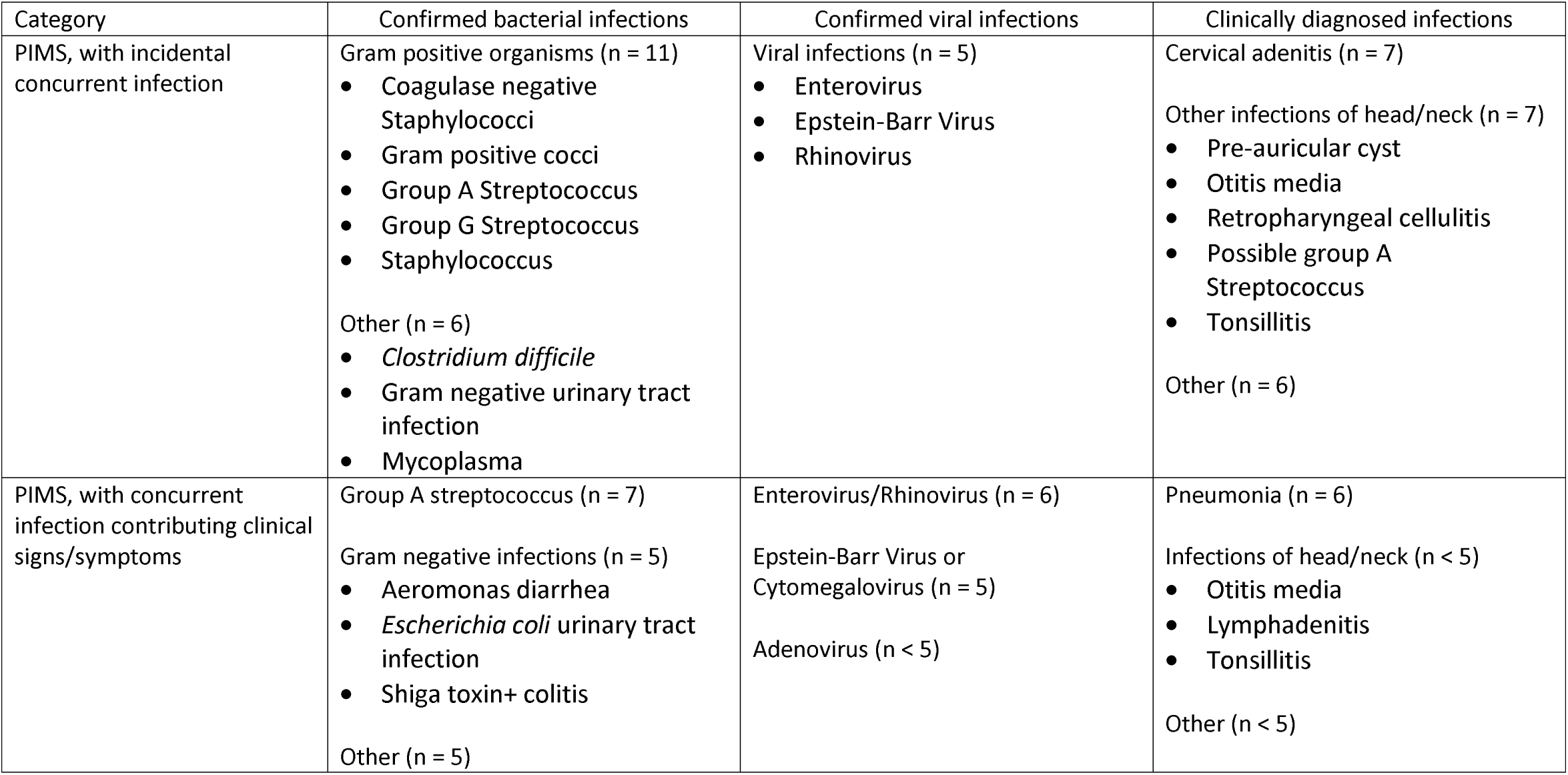
List of reported infections reviewed by investigators, to determine if they may have contributed to some clinical findings.

| Category | Confirmed bacterial infections | Confirmed viral infections | Clinically diagnosed infections |
| --- | --- | --- | --- |
| PIMS, with incidental concurrent infection | Gram positive organisms (n = 11)<br>• Coagulase negative Staphylococci<br>• Gram positive cocci<br>• Group A Streptococcus<br>• Group G Streptococcus<br>• Staphylococcus<br>Other (n = 6)<br>• *Clostridium difficile*<br>• Gram negative urinary tract infection<br>• Mycoplasma | Viral infections (n = 5)<br>• Enterovirus<br>• Epstein-Barr Virus<br>• Rhinovirus | Cervical adenitis (n = 7)<br>Other infections of head/neck (n = 7)<br>• Pre-auricular cyst<br>• Otitis media<br>• Retropharyngeal cellulitis<br>• Possible group A Streptococcus<br>• Tonsillitis<br>Other (n = 6) |
| PIMS, with concurrent infection contributing clinical signs/symptoms | Group A streptococcus (n = 7)<br>Gram negative infections (n = 5)<br>• Aeromonas diarrhea<br>• *Escherichia coli* urinary tract infection<br>• Shiga toxin+ colitis<br>Other (n = 5) | Enterovirus/Rhinovirus (n = 6)<br>Epstein-Barr Virus or Cytomegalovirus (n = 5)<br>Adenovirus (n < 5) | Pneumonia (n = 6)<br>Infections of head/neck (n < 5)<br>• Otitis media<br>• Lymphadenitis<br>• Tonsillitis<br>Other (n < 5) |

**Supplementary Table 7.** Clinical, laboratory, and radiologic features of PIMS hospitalizations by level of care required.

| Characteristics | Level of care required |  | P value |
| --- | --- | --- | --- |
|  | Inpatient ward | Intensive care |  |
| **Number of PIMS hospitalizations, N** | 279 | 127 | --- |
| **Duration of fever (days), median (IQR)<sup>1</sup>** | 7 (5–9) | 6 (5–7) | 0.008 |
| **Clinical features, n (%)** |  |  |  |
| Rash | 205 (73.5) | 83 (65.4) | 0.09 |
| Abdominal pain/vomiting/diarrhea | 195 (69.9) | 117 (92.1) | <0.001 |
| Bilateral bulbar conjunctival injection without exudate | 195 (69.9) | 91 (71.7) | 0.72 |
| Changes in lips/oral cavity | 189 (67.7) | 73 (57.5) | 0.045 |
| Changes in peripheral extremities | 157 (56.3) | 62 (48.8) | 0.16 |
| Cervical lymphadenopathy >1.5 cm diameter | 80 (28.7) | 23 (18.1) | 0.02 |
| Shock/hypotension | 53 (19.0) | 106 (83.5) | <0.001 |
| Periungual desquamation | 49 (17.6) | 14 (11.0) | 0.09 |
| **Coagulation dysfunction, n (%)** | 110 (39.4) | 98 (77.2) | <0.001 |
| **Laboratory features, median (IQR)** |  |  |  |
| Albumin (nadir; $\mu\text{mol/L}$) | 29 (25–35) | 23 (19–28) | <0.001 |
| ALT (peak; U/L) | 30 (17–80) | 54 (32–100) | 0.08 |
| AST (peak; U/L) | 46 (34–80) | 55 (38–100) | 0.27 |
| Bilirubin (peak; $\mu\text{mol/L}$) | 7.0 (4.0–10.1) | 9.7 (6.0–17.8) | 0.20 |
| CRP (peak; mg/L) | 122 (85–199) | 201 (145–260) | <0.001 |
| D-dimer (peak; $\mu\text{g/mL}$) | 1.9 (1.1–4.0) | 3.2 (1.7–4.5) | 0.006 |
| ESR (peak; mm/h) | 56 (43–82) | 51 (36–62) | 0.11 |
| Ferritin (peak; $\mu\text{g/L}$) | 228 (137–433) | 651 (397–1519) | <0.001 |
| LDH (peak; U/L) | 531 (333–833) | 398 (298–686) | 0.03 |
| Platelet count (nadir during admission; $\times 10^9/\text{L}$) | 267 (165–355) | 138 (91–194) | <0.001 |
| Platelet count (peak after admission; $\times 10^9/\text{L}$) | 528 (401–698) | 423 (281–638) | 0.01 |
| Sodium (nadir; $\mu\text{mol/L}$) | 135 (133–137) | 132 (130–135) | <0.001 |
| Troponin (peak; ng/L) | 10 (7–18) | 80 (28–369) | <0.001 |
| **Elevated laboratory features, n / patients with tests (%)** |  |  |  |
| ALT > Institutional 'normal' reference | 127/254 (50.0) | 83/115 (72.2) | <0.001 |
| AST > Institutional 'normal' reference | 73/180 (40.6) | 57/90 (63.3) | <0.001 |
| Bilirubin > Institutional 'normal' reference | 18/104 (17.3) | 19/70 (27.1) | 0.12 |
| CRP $\geq 30$ mg/L | 270/278 (97.1) | 126/127 (99.2) | 0.28 |
| D-dimer > Institutional 'normal' reference | 204/216 (94.4) | 113/115 (98.3) | 0.15 |
| ESR $\geq 40$ mm/h | 176/221 (79.6) | 60/86 (69.8) | 0.07 |
| Ferritin $\geq 500$ $\mu\text{g/L}$ | 52/257 (20.2) | 80/124 (64.5) | <0.001 |
| LDH > Institutional 'normal' reference | 100/203 (49.3) | 62/106 (58.5) | 0.12 |
| Troponin > Institutional 'normal' reference | 53/215 (24.7) | 98/125 (78.4) | <0.001 |
| **Abnormal echocardiogram, n / patients with echo (%)<sup>2</sup>** | 98/266 (36.8) | 93/126 (73.8) | <0.001 |
| Coronary artery lesions, Z-score $\geq 2.5$ | 49/266 (18.4) | 17/126 (13.5) | 0.223 |
| Coronary artery lesions, Z-score < 2.5 | 39/266 (14.7) | 17/126 (13.5) | 0.757 |
| Pericardial effusion | 17/266 (6.4) | 26/126 (20.6) | <0.001 |
| Decreased heart function | 12/266 (4.5) | 59/126 (46.8) | <0.001 |
| Valvular insufficiency | 8/266 (3.0) | 25/126 (19.8) | <0.001 |
| Myocarditis | 7/266 (2.6) | 33/126 (26.2) | <0.001 |
| **Radiologic findings, n / patients with tests (%)** |  |  |  |
| Abnormal chest x-ray | 37/165 (22.4) | 56/117 (47.9) | <0.001 |
| Abnormal ultrasound | 26/70 (37.1) | 22/44 (50.0) | 0.18 |

ALT = Alanine aminotransferase test; AST = Aspartate aminotransferase test; CRP = C-reactive protein; ESR = Erythrocyte sedimentation rate; LDH = Lactate dehydrogenase.
<sup>1</sup>Data available for 386 children (272 inpatient ward; 114 intensive care).
<sup>2</sup>Patients may have multiple cardiac findings, therefore categories exceed total.

**Supplementary Table 8.**
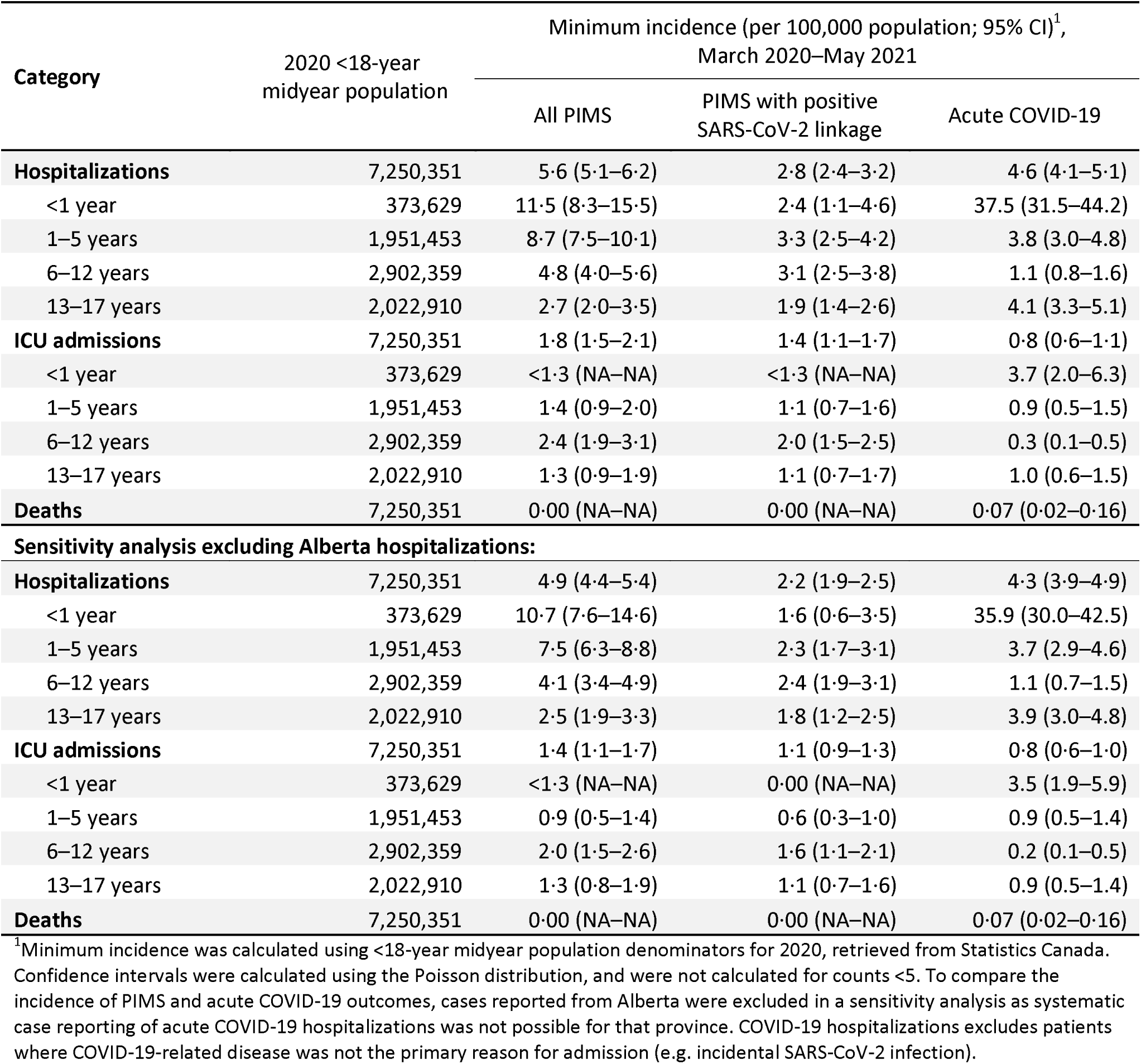
Minimum incidence of PIMS and COVID-19 hospitalization reported to CPSP up until May 2021.

**Supplementary Figure 1.**
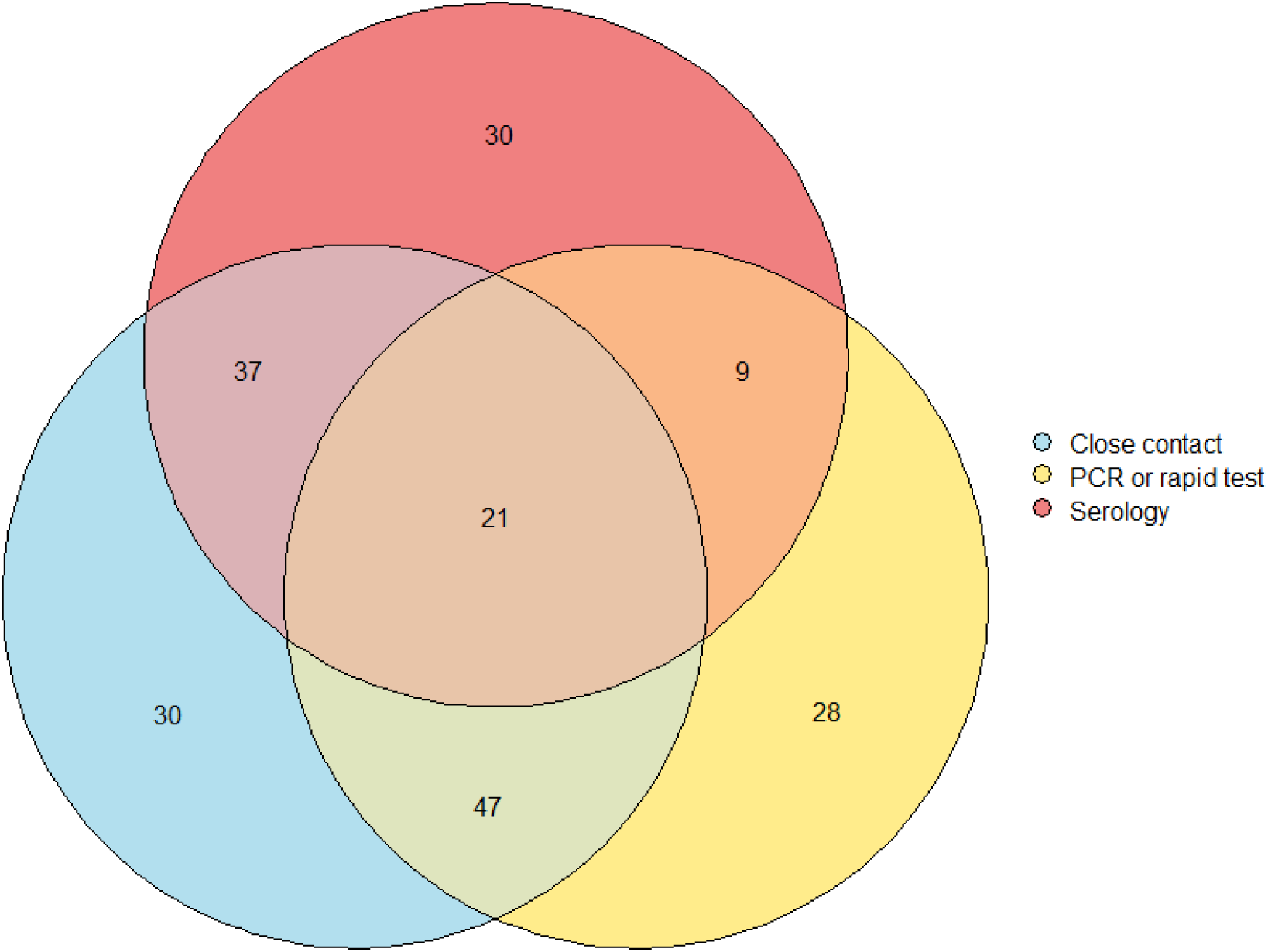
Overlap in epidemiologic links among all 202 PIMS hospitalizations with SARS-CoV-2 exposures, including close contacts with microbiologically confirmed SARS-CoV-2 infection, positive PCR or rapid tests, and positive serology testing.

## Appendix 1: Post-hoc application of the WHO definition for multisystem inflammatory syndrome in children (MIS-C)

### Case definition for MIS-C

In recognition of the adoption of the term “multisystem inflammatory syndrome in children” (MIS-C) by organizations such as the World Health Organization and Centers for Disease Control and Prevention, we applied an adapted post-hoc case definition for MIS-C to all cases of suspected PIMS reported to the CPSP. Given subtle differences in the case definitions for these two related entities, this post-hoc definition was applied regardless of whether cases had previously met the study case definition for PIMS. Cases were classified as MIS-C if the met all of the following criteria (with further detail described in Appendix Table 1):

1. Persistent fever (>38.0°C for ≥ 3 days)
2. Elevated inflammatory markers (CRP ≥ 30 mg/L, ESR ≥ 40 mm/h, and/or ferritin ≥ 500 μg/L)
3. At least two of the following clinical features:
  a. Features of Kawasaki disease
  b. Hypotension or shock
  c. Cardiac involvement
  d. Coagulopathy
  e. Acute gastrointestinal problems
4. Exclusion of alternative etiologies
5. Presence of a SARS-CoV-2 linkage

Criteria 1, 2, and 4 were identical to criteria used in the study case definition for PIMS. Unlike the case definition for PIMS, MIS-C includes evidence of coagulopathy (sub-criterion 3d) and required a confirmed SARS-CoV-2 linkage (criterion 5). In addition, features of Kawasaki disease and toxic shock syndrome were weighted differently for MIS-C than PIMS. Specifically, patients with acute gastrointestinal problems, cardiac involvement, and hypotension/shock more easily met the case definition for MIS-C given a heavier emphasis on other features of Kawasaki disease in the PIMS definition (e.g. bilateral non-purulent conjunctivitis, or mucocutaneous inflammation signs, or rash). The MIS-C definition also did not consider periungual desquamation or cervical lymphadenopathy >1.5 centimetres in diameter as qualifying features of Kawasaki disease, nor elevated liver enzymes as qualifying features of toxic shock syndrome.

#### Overlap of cases meeting MIS-C versus PIMS case definitions

Overall, 209 children reported to the study met the post-hoc case definition for MIS-C. Most cases (n=198, 94.7%) simultaneously met the study case definition for PIMS, though 11 cases (5.3%) met the definition of MIS-C but not PIMS (Appendix Figure 1). All 11 of these patients had reported coagulation dysfunction or elevated D-dimer markers, and lacked a sufficient number of KD-like or TSS-like features and therefore did not meet PIMS criteria. There were 208 cases which met the definition of PIMS but not MIS-C, including <5 cases which met the definition of PIMS with a positive SARS-CoV-2 linkage. Therefore, the analyses of PIMS with a positive linkage (i.e. as presented in the main text) is likely synonymous with results for MIS-C.

#### Characteristics of children hospitalized with MIS-C

The median age at admission for cases of MIS-C was 8· 1 years (IQR 4· 2–11· 5; Appendix Table 2), nearly identical to that of PIMS with a positive SARS-CoV-2 linkage (i.e. median 8· 1 years, IQR 4· 2–11· 9 years). Most cases (n=188, 90.0%) were hospitalized in the second and third waves of the pandemic (i.e. September 2020 onwards), likely due to wider availability of serological testing in Canada to confirm a SARS-CoV-2 linkage as the pandemic progressed. SARS-CoV-2 linkages were most commonly through close contacts with microbiologically-confirmed SARS-CoV-2 infection (n=138, 66.0%), followed by positive PCR testing (n=111, 53.1%), and positive serology (n=98, 46.9%). As with PIMS cases with a positive SARS-CoV-2 linkage, the most common presenting symptom among children with MIS-C was acute gastrointestinal problems (n=189, 90.4%) and a majority of children (n=123, 58.9%) experienced hypotension/shock as well as an abnormal echocardiogram (n=116 out of 204 with echocardiograms performed, 56.9%; Appendix Table 3). Nearly half of children were admitted to intensive care (n=103) as well as required some form of respiratory or hemodynamic support (n=100). No children hospitalized with MIS-C died during the study period.

#### Interpretation of the MIS-C case definition

As confirmed through our post-hoc application of the MIS-C case definition, our findings regarding children with MIS-C and PIMS with a positive SARS-CoV-2 linkage were effectively the same. The primary reason for cases meeting the PIMS but not MIS-C case definition was the lack of a positive SARS-CoV-2 linkage, and may be attributed in part to the lack of widespread serologic testing and other testing modalities in Canada during the early pandemic. Cases of MIS-C and PIMS with a positive SARS-CoV-2 linkage more commonly presented on the severe end of the disease spectrum, with roughly half of children admitted to the ICU and requiring respiratory/hemodynamic support.

We also validated the representativeness of the CPSP reporting system, by comparing the number of cases meeting the post-hoc definition of MIS-C to cases of MIS-C reported to Canada’s Discharge Abstract Database (DAD). The DAD is a national administrative database in Canada where information on all hospital discharges are reported, in all Canadian provinces and territories except Quebec. Cases of MIS-C were identified in the DAD using an ICD-10 diagnostic code of U07.3, with duplicates removed from all counts. Counts from the DAD should be interpreted as provisional numbers, and may not match those published by other sources due to (jurisdictional) variability in data collection and reporting.

Overall, the CPSP ascertained 143 cases of MIS-C outside Quebec, compared to 182 cases of MIS-C outside Quebec from the DAD. We therefore estimate the CPSP reporting system captured roughly 78.6% of MIS-C cases outside Quebec. Notably, the proportion of cases ascertained compared to the DAD fell as the study progressed (Appendix Figure 2). However, given this study did not require a confirmed SARS-CoV-2 linkage, this study identified a large number of cases during the first pandemic wave during which time there was limited serologic testing available in Canada, and which may otherwise have met the MIS-C case definition.

**Appendix Table 1:** Case classification algorithms applied to all cases reported to the CPSP PIMS study.

| Criterion | Paediatric inflammatory multisystem syndrome (PIMS) | Multisystem inflammatory syndrome in children (MIS-C) |
| --- | --- | --- |
| Age of child <18 years | Required | Required |
| Fever (>38° Celsius) of three or more days | Required | Required |
| Evidence of hyperinflammation<br>• C-reactive protein $\geq 30$ mg/L **OR**<br>• Erythrocyte sedimentation rate $\geq 40$ mm/h **OR**<br>• Ferritin $\geq 500$ $\mu\text{g/L}$ | Required | Required |
| PIMS clinical features (**one or both**)<br>• Features of Kawasaki Disease<sup>1</sup><br>• Features of toxic shock syndrome<sup>2</sup> | Required | Not applicable |
| MIS-C clinical features (**at least two**)<br>• Rash, bilateral non-purulent conjunctivitis, or mucocutaneous inflammation signs (oral, hands, or feet)<br>• Hypotension or shock<br>• Cardiac involvement (coronary artery lesions, decreased heart function, myocarditis, valvular insufficiency, or elevated troponin<sup>3</sup>)<br>• Coagulopathy or elevated D-dimer<sup>3</sup><br>• Acute gastrointestinal problems (diarrhea, vomiting, or abdominal pain) | Not applicable | Required |
| Exclusion of alternative aetiologies<sup>4</sup> | Required | Required |
| SARS-CoV-2 linkage<br>• Positive real time polymerase chain reaction **OR**<br>• Positive rapid antigen test **OR**<br>• Positive serologic test **OR**<br>• Close contact with a microbiologically confirmed case of SARS-CoV-2 | Not required | Required |

<sup>1</sup>Features of Kawasaki Disease defined as two or more of the following: rash, bilateral non-purulent conjunctivitis, mucocutaneous inflammation signs (oral, hands, or feet), periungual desquamation, cervical lymphadenopathy >1.5 centimetres in diameter, or cardiac involvement (including any of coronary artery lesions, decreased heart function, myocarditis, valvular insufficiency, or elevated troponin). The presence of cardiac involvement only also satisfies this criterion.
<sup>2</sup>Features of toxic shock syndrome defined as: a) at least one of rash, periungual desquamation, or hypotension/shock; plus b) at least one of abdominal pain/vomiting/diarrhea, bilateral non-purulent conjunctivitis, changes in the lips or oral cavity, or elevated liver enzymes (i.e. ALT, AST, LDH, or bilirubin; determination of elevated markers below).
<sup>3</sup>To assess laboratory criteria, investigators were asked to report their institutional age- and sex-specific laboratory reference ranges for D-dimer, liver enzymes, and troponin. Where references were not provided (24.4% of cases), references from the Canadian Laboratory Initiative on Paediatric Reference Intervals (CALIPER) study were used, while an assumed threshold of 0.5 $\mu\text{g/mL}$ was used for D-dimer\*. Patient markers above their corresponding institutional or CALIPER reference range were categorized as elevated.
<sup>4</sup>Determined by dual physician review of cases with concurrent microbiologically confirmed or clinically diagnosed infections, and consensus discussion.
\*Adeli, Khosrow et al. "The Canadian laboratory initiative on paediatric reference intervals: A CALIPER white paper." *Critical reviews in clinical laboratory sciences* vol. 54,6 (2017): 358-413. doi:10.1080/10408363.2017.1379945

**Appendix Table 2.** Demographic and outcome characteristics of children who met the WHO definition of MIS-C.

| **Characteristics, n (%)** | **Distribution** |
| --- | --- |
| **Number of MIS-C hospitalizations, N** | 209 |
| **Age (years), median (IQR)** | 8.1 (4.2–11.5) |
| **Age of child (years), n (%)** |  |
| Infants (<1 year) | 9 (4.3) |
| Preschool (1-5 years) | 67 (32.1) |
| School age (6-12 years) | 93 (44.5) |
| Adolescents (13-17 years) | 40 (19.1) |
| **Sex of child, n (%)** |  |
| Female | 76 (36.4) |
| Male | 133 (63.6) |
| **Positive SARS-CoV-2 linkage, n (%)<sup>1</sup>** | 209 (100.0) |
| Close contact with confirmed SARS-CoV-2 | 138 (66.0) |
| PCR positive | 111 (53.1) |
| Serology positive | 98 (46.9) |
| **Phase of pandemic, n (%)** |  |
| Wave 1 (Mar–Aug 2020) | 21 (10.0) |
| Wave 2/3 (Sep 2020–May 2021) | 188 (90.0) |
| **Admitted to ICU, n (%)** | 103 (49.3) |
| **Respiratory/hemodynamic support required, n (%)** | 100 (47.8) |
| **Child died, n (%)** | 0 (0.0) |

<sup>1</sup>Defined as a positive PCR or rapid test, serology, or close contact.

**Appendix Table 3.** Clinical, laboratory, and radiologic features of children who met the WHO definition of MIS-C.

| Characteristics | Distribution |
| --- | --- |
| **Number of MIS-C hospitalizations, N** | 209 |
| **Duration of fever (days), median (IQR)<sup>1</sup>** | 6 (5–8) |
| **Clinical features, n (%)** |  |
| Abdominal pain/vomiting/diarrhea | 189 (90.4) |
| Bilateral bulbar conjunctival injection without exudate | 146 (69.9) |
| Rash | 135 (64.6) |
| Shock/hypotension | 123 (58.9) |
| Changes in lips/oral cavity | 113 (54.1) |
| Changes in peripheral extremities | 88 (42.1) |
| Cervical lymphadenopathy >1.5 cm diameter | 35 (16.7) |
| Periungual desquamation | 16 (7.7) |
| **Coagulation dysfunction, n (%)** | 140 (67.0) |
| **Laboratory features, median (IQR)** |  |
| Albumin (nadir; $\mu\text{mol/L}$) | 25 (20–29) |
| ALT (peak; U/L) | 45 (22–91) |
| AST (peak; U/L) | 50 (37–95) |
| Bilirubin (peak; $\mu\text{mol/L}$) | 9.5 (6.0–18.1) |
| CRP (peak; mg/L) | 181 (120–239) |
| D-dimer (peak; $\mu\text{g/mL}$) | 2.4 (1.5–4.2) |
| ESR (peak; mm/h) | 51 (38–70) |
| Ferritin (peak; $\mu\text{g/L}$) | 470 (277–970) |
| LDH (peak; U/L) | 407 (299–722) |
| Platelet count (nadir during admission; $\times 10^9/\text{L}$) | 149 (112–212) |
| Platelet count (peak after admission; $\times 10^9/\text{L}$) | 446 (327–603) |
| Sodium (nadir; $\mu\text{mol/L}$) | 133 (130–136) |
| Troponin (peak; ng/L) | 47 (10–199) |
| **Elevated laboratory features<sup>2</sup>, n / patients with tests (%)** |  |
| ALT > Institutional 'normal' reference | 111/184 (60.3) |
| AST > Institutional 'normal' reference | 71/141 (50.4) |
| Bilirubin > Institutional 'normal' reference | 26/103 (25.2) |
| CRP $\geq 30$ mg/L | 207/208 (99.5) |
| D-dimer > Institutional 'normal' reference | 187/191 (97.9) |
| ESR $\geq 40$ mm/h | 105/146 (71.9) |
| Ferritin $\geq 500$ $\mu\text{g/L}$ | 98/208 (47.1) |
| LDH > Institutional 'normal' reference | 85/167 (50.9) |
| Troponin > Institutional 'normal' reference | 123/200 (61.5) |
| **Abnormal echocardiogram, n / patients with echo (%)<sup>2</sup>** | 116/204 (56.9) |
| Decreased heart function | 63/204 (30.9) |
| Myocarditis | 35/204 (17.2) |
| Pericardial effusion | 30/204 (14.7) |
| Coronary artery lesions, Z-score $\geq 2.5$ | 27/204 (13.2) |
| Coronary artery lesions, Z-score < 2.5 | 27/204 (13.2) |
| Valvular insufficiency | 25/204 (12.3) |
| **Radiologic findings, n / patients with imaging (%)** |  |
| Abnormal chest x-ray | 69/171 (40.4) |
| Abnormal CT scan | 17/27 (63.0) |
| Abnormal MRI scan | 5/13 (38.5) |
| Abnormal ultrasound | 29/57 (50.9) |

ALT = Alanine aminotransferase test; AST = Aspartate aminotransferase test; CRP = C-reactive protein; CT = Computed tomography; ESR = Erythrocyte sedimentation rate; LDH = Lactate dehydrogenase; MRI = Magnetic resonance imaging.
<sup>1</sup>Data available for 195 cases.
<sup>2</sup>Patients may have multiple cardiac findings, therefore categories exceed total.

**Appendix Figure 1.**
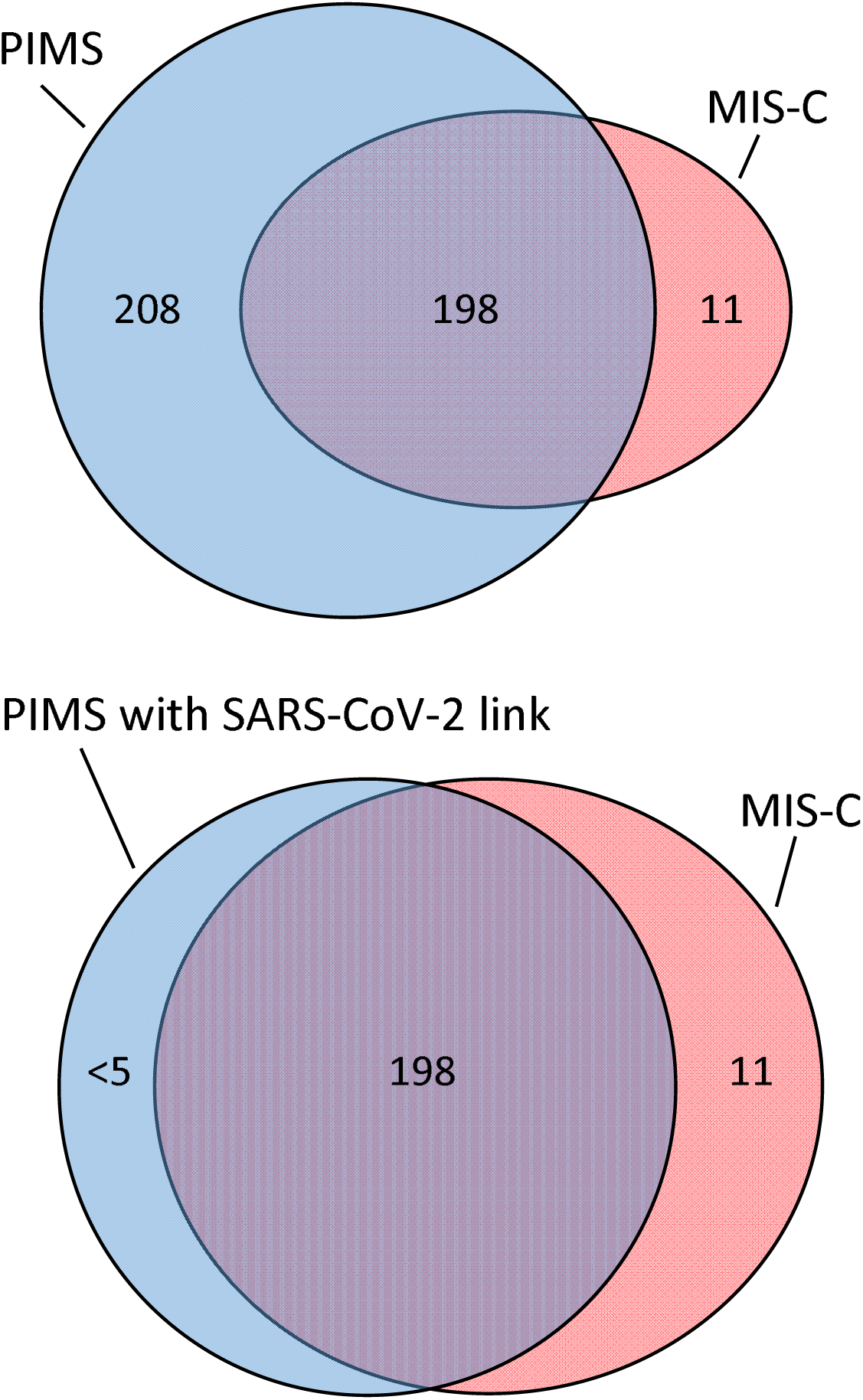
Overlap of CPSP PIMS surveillance definition and post-hoc application of WHO definition of multisystem inflammatory syndrome in children (MIS-C).

Eleven patients met criteria for MIS-C but not PIMS due to variation in the definitions applied (see Appendix 1). All 11 patients had reported coagulation dysfunction or elevated D-dimer markers, which is considered in the MIS-C but not the PIMS definition. In addition, these cases lacked a sufficient number of KD-like features (i.e. two required to meet PIMS definition) or TSS-like features (i.e. one from each of two groups of symptoms to meet PIMS definition) and therefore did not meet PIMS criteria.

**Appendix Figure 2.**
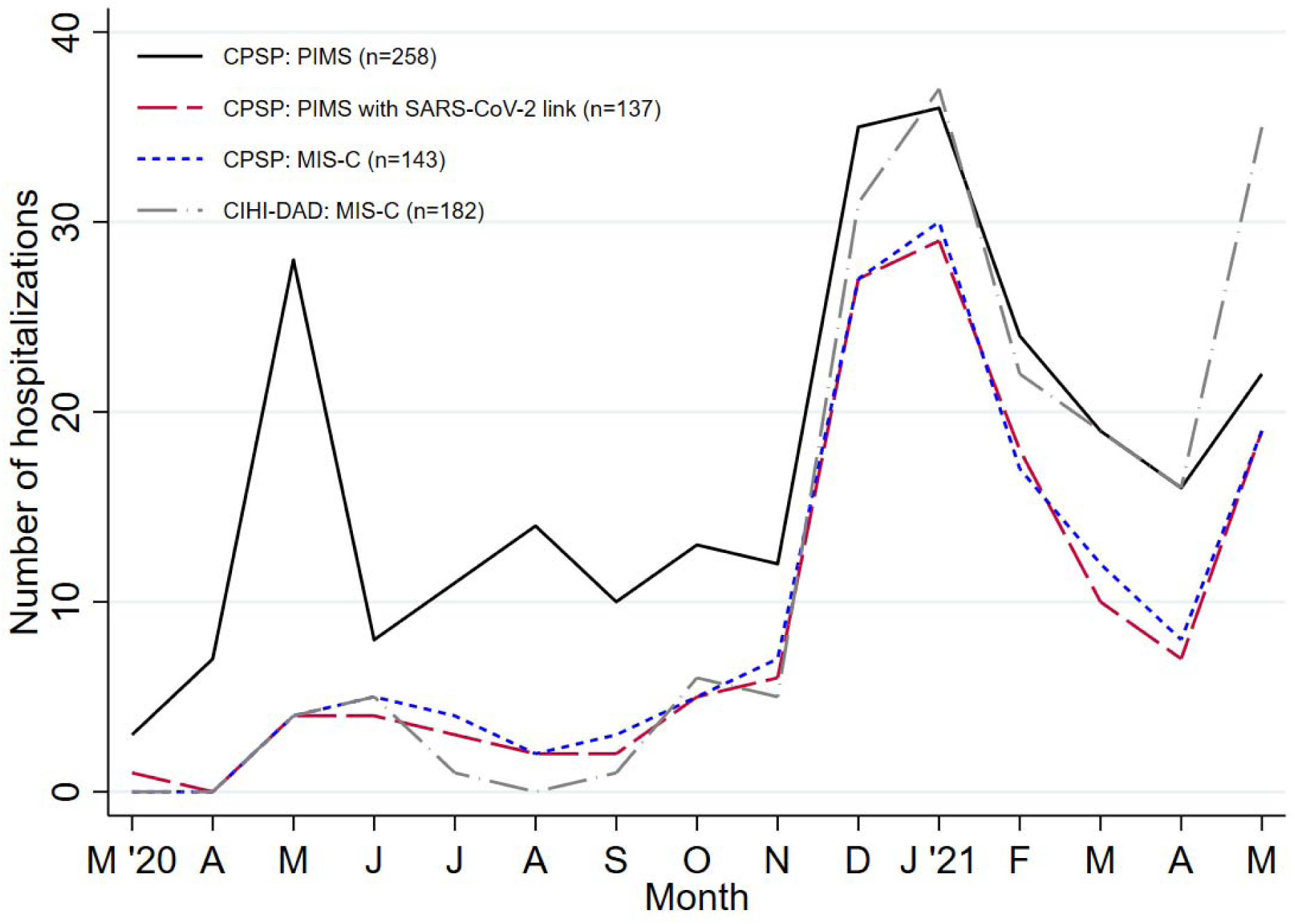
Number of hospitalizations per month using the CPSP PIMS study definition, CPSP PIMS with SARS-CoV-2 linkage definition, CPSP post-hoc MIS-C definition, and MIS-C cases from the Canadian Institute for Health Information’s Discharge Abstract Database (DAD).

Data from the DAD depict children and youth aged 0-17 (inclusive) admitted to hospital with an ICD-10 diagnostic code of U07.3 (Multisystem Inflammatory Syndrome). Duplicate cases were identified using a unique identifier and then removed from the counts. Quebec data are not included as they are not available in the DAD, and therefore CPSP cases from Quebec were also excluded from this figure. Data from the DAD are provisional, may be incomplete or subject to change, and counts may not match those published by other sources due to (jurisdictional) variability in data collection and reporting.

